# Accessory Genomes Drive Independent Spread of Carbapenem-Resistant *Klebsiella pneumoniae* Clonal Groups 258 and 307

**DOI:** 10.1101/2021.08.04.21261380

**Authors:** William C Shropshire, An Q Dinh, Michelle Earley, Lauren Komarow, Diana Panesso, Kirsten Rydell, Sara I Gómez-Villegas, Hongyu Miao, Carol Hill, Liang Chen, Robin Patel, Bettina C Fries, Lilian Abbo, Eric Cober, Sara Revolinski, Courtney L Luterbach, Henry Chambers, Vance G Fowler, Robert A Bonomo, Samuel A Shelburne, Barry N Kreiswirth, David van Duin, Blake M Hanson, Cesar A Arias

**Affiliations:** Center for Infectious Diseases, School of Public Health, University of Texas Health Science Center, Houston, Texas, 77030 USA; Center for Antimicrobial Resistance and Microbial Genomics, Division of Infectious Diseases, University of Texas Health Science Center at Houston McGovern Medical School, Houston, Texas, 77030 USA; The Biostatistics Center, The George Washington University, Rockville, MD, USA; Department of Microbiology and Molecular Genetics, University of Texas McGovern Medical School at Houston, Houston, Texas, 77030 USA; Molecular Genetics and Antimicrobial Resistance Unit-International Center for Microbial Genomics, Universidad El Bosque, Bogotá, Colombia; Department of Biostatistics and Data Science, School of Public Health, University of Texas Health Science Center, Houston, Texas, 77030 USA; Duke Clinical Research Institute, Duke University Medical Center, Durham, NC, USA; Center for Discovery and Innovation, Hackensack Meridian Health, Nutley, NJ, USA; Division of Clinical Microbiology, Department of Laboratory Medicine and Pathology, and Division of Infectious Diseases, Department of Medicine, Mayo Clinic, Rochester, MN, USA; Department of Medicine, Infectious Disease Division, Stony Brook University, Stony Brook, NY, USA; Veteran’s Administration Medical Center, Northport, New York, USA; Division of Infectious Diseases, Department of Medicine, University of Miami Miller School of Medicine and Jackson Health System, Miami, FL, USA; Department of Infectious Diseases, Cleveland Clinic, Cleveland, OH, USA; School of Pharmacy, Medical College of Wisconsin, Milwaukee, WI, USA; Division of Infectious Diseases, University of North Carolina at Chapel Hill, Raleigh, NC, USA; Department of Medicine, University of California San Francisco, San Francisco, USA; Division of Infectious Diseases, Duke University, Durham, NC, USA; Departments of Pharmacology, Molecular Biology and Microbiology, Biochemistry, and Proteomics and Bioinformatics, Case Western Reserve University School of Medicine, Cleveland, OH, USA; CWRU-Cleveland VAMC Center for Antimicrobial Resistance and Epidemiology, Cleveland, OH, USA; Department of Infectious Diseases, The University of Texas MD Anderson Cancer Center, Houston, Texas, USA; Department of Genomic Medicine, The University of Texas MD Anderson Cancer Center, Houston, Texas, USA

**Keywords:** Carbapenem-resistant *Klebsiella pneumoniae*, mobile genetic elements, genomic surveillance, divergent evolution

## Abstract

**Background:** Carbapenem-resistant *Klebsiella pneumoniae* (CR*Kp*) are urgent public health threats. Worldwide dissemination of CR*Kp* has been largely attributed to the clonal group (CG) 258. However, recent evidence indicates the global emergence of a CR*Kp* CG307 lineage. Houston, Texas is the first large city in the US with co-circulation of both CR*Kp* CG307 and CG258. We sought to characterize the genomic and clinical factors contributing to the parallel endemic spread of CG258 and CG307.

**Methods:** CR*Kp* isolates were collected as part of the prospective, Consortium on Resistance Against Carbapenems in *Klebsiella* and other *Enterobacterales* 2 (CRACKLE-2) study. Hybrid short-read and long-read genome assemblies were generated from 119 CR*Kp* isolates (95 originated from Houston hospitals). A comprehensive characterization of phylogenies, gene transfer, and plasmid content with pan-genome analysis were performed on all CR*Kp* isolates. Plasmid mating experiments were performed with CG307 and CG258 isolates of interest. An inverse-probability weighted Desirability of Ordinal Outcome Ranking (DOOR) analysis was conducted to determine if patients infected/colonized with CG307 had differences in overall clinical outcomes from patients infected/colonized with CG258.

**Results:** Dissection of the accessory genomes suggested independent evolution and limited horizontal gene transfer between CG307 and CG258 lineages. CG307 contained a diverse repertoire of mobile genetic elements harboring carbapenemases, which were shared with other non-CG258 *K. pneumoniae* isolates. Three unique clades of Houston CG307 isolates contained a diverse repertoire of mobile genetic elements harboring carbapenemases and clustered distinctly from other global CG307 isolates. CG307 were often isolated from the urine of hospitalized patients, likely serving as important reservoirs for genes encoding carbapenemases and extended-spectrum beta-lactamases. The DOOR probability estimate (64%; 95% CI: 48, 79) of our Houston-based cohort suggested that there was a general trend for patients infected/colonized with CG307 to have more favorable outcomes than patients infected/colonized with CG258.

**Conclusions:** Our findings suggest parallel co-circulation of high-risk lineages with potentially divergent evolution. CG307 is widely circulating CR*Kp* clone in the Houston region with the potential to transfer major resistance determinants to other non-CG258 CR*Kp* lineages. Our findings provide major insights into the mechanism of epidemic spread of CR*Kp*.

## Background

Carbapenem-resistant *Klebsiella pneumoniae* (CR*Kp*) cause significant worldwide morbidity and mortality and are of paramount concern in nosocomial settings (1). Since the identification of *K. pneumoniae* isolates harboring the carbapenem-hydrolyzing enzyme *K. pneumoniae* carbapenemase (KPC) in 1996, global dissemination of CR*Kp* has occurred (2). CR*Kp* now make up the majority of carbapenem-resistant *Enterobacterales* (CRE) infections in the United States (3, 4). Infections caused by CR*Kp* are difficult to treat since these isolates often harbor resistance to multiple antibiotics, limiting therapeutic options (5).

Global dissemination of CR*Kp* has been largely attributed to the clonal expansion of a genetic lineage of *K. pneumoniae* designated clonal group 258 (CG258), first identified in the United States in 2008 (6–8). Spread of CG258 has been associated with carriage of genes encoding the KPC enzyme (6, 9) on F-type pKpQIL plasmids (10). More recently, another CR*Kp* genetic lineage, designated CG307, has emerged and appears to be spreading in countries such as Italy, Pakistan, Colombia, and the United States (11–13). Isolates in the *K. pneumoniae* CG307 lineage carry distinct genomic features that may confer virulence and colonization advantages, such as F-type plasmid-borne glycogen synthesis gene clusters and urea transport systems (12). These potential virulence determinants are present in conjunction with established pathogenic factors, such as the *mrk* gene cluster (encoding a type 3 pili), and extracellular polysaccharides (capsule and lipopolysaccharides) found across all MDR *K. pneumoniae* lineages (14). Carriage of *bla*_KPC_ on pKpQIL and N-type plasmids in CG307 isolates has been documented (12); however, not at the same prevalence as CG258. There is a strong association of the CG307 lineage with the carriage of *bla*_CTX-M-15_, a gene encoding an extended-spectrum β-lactamase (ESBL) (11). Of interest, a molecular dating analysis estimated that CG307 arose in 1994 (11) near the estimated year of emergence of CG258 (6), suggesting that these two high-risk genetic lineages have evolved in parallel.

A recent large US cohort study of patients infected or colonized with CRE (Consortium on Resistance Against Carbapenems in *Klebsiella* and other *Enterobacterales* 2 – CRACKLE-2 study) identified *K. pneumoniae* CG307 as the second most common lineage of CR*Kp* after CG258 (7% vs 64% out of 593 isolates, respectively) (3). Of note, in that study, the majority (72%) of the CG307 *K. pneumoniae* isolates were found in the Houston, TX metropolitan region. This observation is in agreement with previous reports describing the molecular epidemiology of multidrug-resistant *K. pneumoniae* in a hospital network in Houston, TX (15). Furthermore, the CRACKLE-2 results suggested that Houston had become the first large city in the United States with high endemicity of CR*Kp* where *K. pneumoniae* CG307 and CG258 appear to be co-circulating and undergoing parallel expansion. It is unclear how these respective lineages were introduced in the same geographical region, share antimicrobial resistance and virulence determinants, and to what extent these clonal groups impact clinical outcomes.

To dissect factors that drove the parallel co-circulation of CG307 and CG258, as well as the high endemicity of CR*Kp* in the Houston area, we generated complete assemblies of *K. pneumoniae sensu stricto* recovered from patients enrolled in the CRACKLE-2 cohort in Houston and other sites in the United States. We assessed potential intra- and inter-clade transmission of vectors responsible for carbapenem resistance between CG258 and CG307, their respective correlated gene content, and their association with clinical outcomes in patients colonized or infected with CR*Kp*.

## Methods

### Study design

Characterization of patients and isolates was based on the CRACKLE-2 study(3), a prospective, multicenter observational cohort study performed in 49 hospitals across the continental United States. This study focused on patients and their isolates recruited in Houston, TX. A total of 160 CRE isolates were collected from 10 hospital sites within the Houston, TX metropolitan area from May 2017 to December 2017. A total of 95/160 CRE (59.4%) were identified as belonging to the *K. pneumoniae* species complex (KpSC) and were subjected to whole genome sequencing. The KpSC is comprised of 7 similar phylogroups (∼98-99% average nucleotide identity) including *K. pneumoniae sensu stricto* (Kp1; n = 94) and *K. pneumoniae quasipneumoniae* subsp. *quasipneumoniae* (Kp2; n = 1), which were both found in our study (5, 16). Additionally, we included all other CG307 isolates identified outside of the Houston metropolitan area (n=12) from the CRACKLE-2 study (3), along with 12 geographically matched patients infected/colonized with CG258 isolates for purposes of comparing genomic and population structure differences.

### Whole genome sequencing

Isolate culture, genomic DNA extraction, short-read sequencing library preparation, and Illumina short-read sequencing have been described previously (3). All isolates identified as CR*Kp* in our sampling frame were subjected to Oxford Nanopore Technologies (ONT) long-read sequencing using the SQK-RBK004 library preparation kit and sequenced on an Oxford Nanopore GridION X5 (Oxford, UK). MinKNOW-v3.0.13 software was used for fast5 generation with subsequent GPU-enabled base-calling and read filtering (minimum length of 1000 bp; Phred score ≥ Q7) performed with Guppy-v3.2.2 software. Raw ONT fastq reads were trimmed and de-multiplexed with qcat-v1.1.0 (nanoporetech, GitHub: https://github.com/nanoporetech/qcat).

### Consensus hybrid bacterial genome assembly pipeline

A customized python script was used for short- and long-read hybrid genome assemblies (Shropshire, W GitHub: https://github.com/wshropshire/flye_hybrid_assembly_pipeline). Briefly, raw assemblies were created with Flye-v2.7 (17). Raw assemblies then underwent a series of error correction steps using first an iterative long-read polish step with Racon-v1.4.5 (18) followed by Medaka-v0.11.5 (nanoporetech GitHub: https://github.com/nanoporetech/medaka). The consensus medaka assembly then underwent a series of Racon short-read polishing steps with a final error correction step using a modified python script (powerpak GitHub: https://github.com/powerpak/pathogendb-pipeline/tree/master/scripts) that fixes regions with low, short-read coverage. Fragmented assemblies were re-assembled with Unicycler-v0.4.6 (19) and compared with Flye assemblies for accuracy and contiguity. Each assembly was manually curated using the Integrative Genomics Viewer (IGV-v2.8.0) to visually inspect contiguity and accuracy of assemblies using both short-read and long-read alignments (20–22). Further quality control with additional preliminary results were generated using a customized python script (Shropshire, W GitHub: https://github.com/wshropshire/QA_QC_tool).

Genome completeness and contamination data were assessed using BUSCO.v4 (23) and CheckM-v1.1.2 (24). Genome completeness, as defined by proportion of lineage specific, colocalized marker gene sets in *Enterobacterales* (24), was greater than 98% for the entire set of 119 KpSC isolates. Genome contamination as defined by multiple copies of these aforementioned marker gene sets (24) was less than 4% for all isolates. These estimates were within the mean absolute error of completeness and contamination using lineage specific sets with all isolates meeting the designation of near complete genomes (24). Assembly metrics are shown in Additional file 1: Table S1 with their respective BioSample Accession and ARLG identification numbers.

### Molecular and genomic characterization

Genome annotation was performed using Prokka-v1.14.5 (25). Capsule typing (26, 27), multi-locus sequence typing (MLST), virulence factor (28, 29), and antimicrobial factor quantification was completed using Kleborate-v1.0.0 (30). Clonal groups were defined as similarly related isolates that may only differ by one of seven MLST housekeeping gene alleles as identified by the Pasteur Institute using their BIGSdb (https://bigsdb.pasteur.fr/klebsiella/klebsiella.html). Kleborate virulence scores are reported from 0 to 5, as follows: 0, negative for yersiniabactin (*ybt*), colibactin *(clb*), and aerobactin (*iuc*); 1, yersiniabactin only; 2, colibactin with or without yersiniabactin; 3, aerobactin only; 4, aerobactin with yersiniabactin; 5, all three virulence determinants. Kleborate resistance scores range from 0 to 3 with the following definitions: 0, no ESBL, no carbapenemase; 1, ESBL, no carbapenemase; 2, carbapenemase positive without colistin resistance; 3, carbapenemase with colistin resistance. The ABRicate-v0.9.8 tool (Seemann, T GitHub: https://github.com/tseemann/abricate) was used in parallel with Kleborate to query the Comprehensive Antibiotic Resistance Database (CARD) (31) and PlasmidFinder (32) databases to characterize antibiotic resistant and plasmid determinants, respectively (Accessed 2020-06-17). A summary of the Kleborate output can be found in Additional file 1: Table S2. A matrix of all *in silico* PCR-based replicon typing found with PlasmidFinder is included in Additional file 1: Table S3.

### Pangenome analyses

Panaroo-v1.2.2 (33) was used to generate a pangenome profile from the 118 gff files generated from Prokka-v1.14.5 (25) using the ‘sensitive’ clean-mode for gene contamination filtering. Three reference isolates were included: NJST258_1 (GenBank Accession #: GCA_000598005.1), NJST258_2 (GenBank Accession #: GCA_000597905.1), and KPN11 (GenBank Accession #: GCA_002148835.1). Isolate C719 was not included in pan-genome analysis since it is a member of the *K. quasipneumoniae* subsp. *quasipneumoniae* (Kp2) phylogroup (34). Clusters of orthologous gene groups present in greater than ≥ our total group were aligned using MAFFT-v7.470 (35) to create a core gene alignment. Snp-dists (Seemann, T snp-dists https://github.com/tseemann/snp-dists) was used to create a pairwise SNP distance matrix from the core gene alignment. Median nucleotide divergence of core genes analyzed was calculated by taking median pairwise SNP differences and dividing by total length of the core gene alignment minus ambiguous characters and INDELs (3,768,044 bp).

IQ-TREE-v2.0.6 (36) was used to create a ML phylogeny inferred tree using the core gene alignment in order to determine population structure. ModelFinder (37) was used to determine the best model fit given our data, which was a GTR substitution model with a rate heterogeneity ‘FreeRate’ model using 3 categories. We performed non-parametric bootstrapping (n = 1,000 replicates) and implemented transfer bootstrap expectation (TBE) (38) to create branch bootstrap support values for our final trees. Hierarchical population structure was investigated using the core gene alignment with rhierBAPS-v1.1.3 (39). Tree visualization output was achieved using iTOL (40).

A Jaccard distance matrix was created using the gene presence/absence (n=13049) matrix from the pan-genome analysis with the CRAN package “Philentropy” (41). The partitioning around medoid (PAM) algorithm was used with varying cluster sizes (k =1, k = 5) to determine cluster assignment using the CRAN package “cluster” (42). We used the PANINI tool (43) that utilizes the unsupervised clustering t-Distributed Stochastic Neighbor Embedding (t-SNE) method with the Barnes-Hut algorithm to determine how the accessory genome clusters into groups. A principal component analysis (PCA) was performed using the base R ‘prcomp()’ function. The pan-genome reference fasta file generated by Panaroo was functionally annotated for the purpose of observing orthology relationships using EggNOG-v.5 (44, 45). CG307 genome comparisons were generated using the CGViewer Comparison Tool (46). Phage content for representative strains were characterized using PHASTER (47, 48). Data visualization for all analysis performed in R was generated with ggplot2 (49).

### Core SNP phylogenies

A core SNP phylogeny was conducted using C268 (CG258) and C246 (CG307) as a reference for short-read alignment using the ‘snippy-multi’ wrapper tool in Snippy-v4.6.0 (Seemann, T GitHub: https://github.com/tseemann/snippy) for Houston-only isolates. Similarly, a core SNP phylogeny with CG307 isolates collected from NCBI was performed using C234 as a reference. For SRA only submissions (15), SRA files were retrieved using sratools-v2.10.9 and assembled using SPAdes-v3.14.0. Quality control on assemblies (*e.g.* ambiguous bases, incorrect genome size, contamination) was performed using Kleborate-v2.0.1. Recombination regions were masked using Gubbins-v2.3.4 (50). Subsequently, the filtered alignment file was used as an input to infer a core SNP, maximum-likelihood phylogeny using IQ-TREE-v2.0.6 (36).

### Plasmidome characterization

*In silico* plasmid typing and characterization was performed using the MOB-suite, MOB-typer tool (51). Details on all 424 extrachromosomal contigs identified using MOB-typer tool, including replication initiation protein, relaxase, mate-pair formation, and host range prediction are included in Additional file 1: Table S4. We created a sketch representation of the approximate plasmid sequence distances using the MinHash technique with the Mash toolkit (52) at both the isolate (n = 119) and individual plasmid level. For plasmid type comparisons, plasmids below 25 kbp, mostly ColE1-like plasmids, were excluded from analysis due to large differences when compared to larger (> 25 kbp) size plasmids. The distance matrices were then used to generate a heatmap and dendrogram using the heatmap.2 (talgalili, GitHub: https://github.com/talgalili/gplots) R package. The dendrogram was created through agglomerative hierarchical clustering using an ‘average’ linkage.

### Conjugation transfer assays

Four isolates with plasmids of interest based on plasmidome analysis were included in conjugation transfer assays using a modified protocol (53). Details for the particular plasmids studied can be found in Additional file 1: Table S5. Easyfig was used to compare plasmid structures (54). Donors and a sodium azide (NaN_3_)-resistant *E. coli* strain J53 were grown up overnight in TSB supplemented with 2 µg/mL ertapenem or 10 µg/mL gentamicin for plasmid selection and 150 µg/mL NaN_3_ for counter-selection at 37°C with mild agitation. Overnight cultures were washed in 0.9% NaCl 2X and then sub-cultured into fresh TSB at a 1:100 dilution and incubated at 37°C until mid-log phase (∼0.6 OD600). Broth mating was performed with 1:10 donor-to-recipient ratios with TSB overnight at 37°C for 20 hours. TSA plates supplemented with 2 µg/mL ertapenem or 10 µg/mL gentamicin and 150 µg/mL NaN_3_ were used to select for transconjugants. Conjugation transfer frequency was enumerated by calculating the ratio of CFU/ml in transconjugants over the CFU/ml in the donor plates respectively. The limit of detection was calculated by taking the minimum CFU threshold detected factoring in dilution factor (50 CFU/mL) and dividing by donor frequency. We used a PCR protocol to check for the positive identification of transconjugants screening for *bla*_KPC-2_, *aacA4, E. coli* J53 (*rpoB* gene), as well as plasmids of interest with primers listed in Additional file 1: Table S6.

### Clinical data outcomes assessed using desirability of ordinal outcome ranking (DOOR) analysis

The Houston-based CG258 (n=27) isolates and CG307 (n=32) isolates were included in an inverse probability weighted (IPW)-adjusted Desirability of Ordinal Outcome Ranking (DOOR) analysis (55) in which composite patient level outcomes were compared. Three detrimental outcomes (no clinical response, unsuccessful discharge, adverse event) were used to create the ordinal DOOR outcome assessed 30-days after culture, as described before (3). Briefly, the outcomes were defined as follows: (1) ‘clinical response’ was an improvement in symptoms with no additional antibiotic course that has *in vitro* activity against first positive culture and no subsequent relapse; (2) ‘unsuccessful discharge’ was post-culture stay ≥ 30 days or readmission within 30 days; (3) patients had an ‘adverse event’ if subsequent *Clostridioides difficile* infection or post-culture renal failure occurred. The most favorable outcome was ‘(1) alive without events’ whereas the worst outcome was (4) death; intermediate levels were (2): alive with one event and (3): alive with 2 or 3 events. An IPW-adjusted DOOR probability estimate of composite outcomes between patients in each of the two clonal groups was assessed, with a 50% probability estimate indicating comparable outcomes. A DOOR probability estimate greater than 50% with a 95% confidence interval above 50% indicates, with statistical significance, a more favorable outcome for one randomly selected member of a group compared to a randomly selected member of another group. Covariates that were included into the weights were age, Charlson comorbidity index, and origin of patient admission. Non-parametric bootstrap methods (n=4000) were used to estimate 95% confidence intervals.

### Statistics

Group level distributions of continuous variables were evaluated using one-way ANOVA test or Kruskal-Wallis rank-sum tests contingent on assumptions based on the data distributions. *Post-hoc* tests for continuous variables with significant ANOVA p-values were accomplished using the Tukey’s honest significance test or the Dunn test using the “FDR” method to control for multiple comparisons. Wilcoxon rank-sum test was used to determine distribution differences of continuous variables across two groups. Distributions of categorical data were evaluated using Pearson Chi Square or Fisher’s Exact tests, dependent on cell counts. All statistical analysis was performed with R-v4.0.0 software.

## Results

### Molecular epidemiological features of CG258 and CG307 circulating in same geographical area reveal distinct virulence and antimicrobial resistance gene content

The majority of isolates identified as CR*Kp* belonged to the taxa *K. pneumoniae sensu stricto* (94/95; 98.9%), with the exception of one isolate (C719, identified as *K. quasipneumoniae* subsp. *quasipneumoniae*). The predominant CR*Kp* sequence types identified in the Houston cohort were CG258 (37/95; 38.9%) and CG307 (35/95; 36.8%). The remaining isolates belonged to CG15 (n=5), CG20 (n=4), CG147 (n=4) and a mixture of other sequence types (n = 10) (Fig. 1). There were no observed correlations between clonal group and hospital location across the Houston metropolitan area (p = 0.8), suggesting each site had a similar distribution to the overall circulation of sequence types in this hospital system (Additional file 2: Fig. S1).

**Fig. 1.**
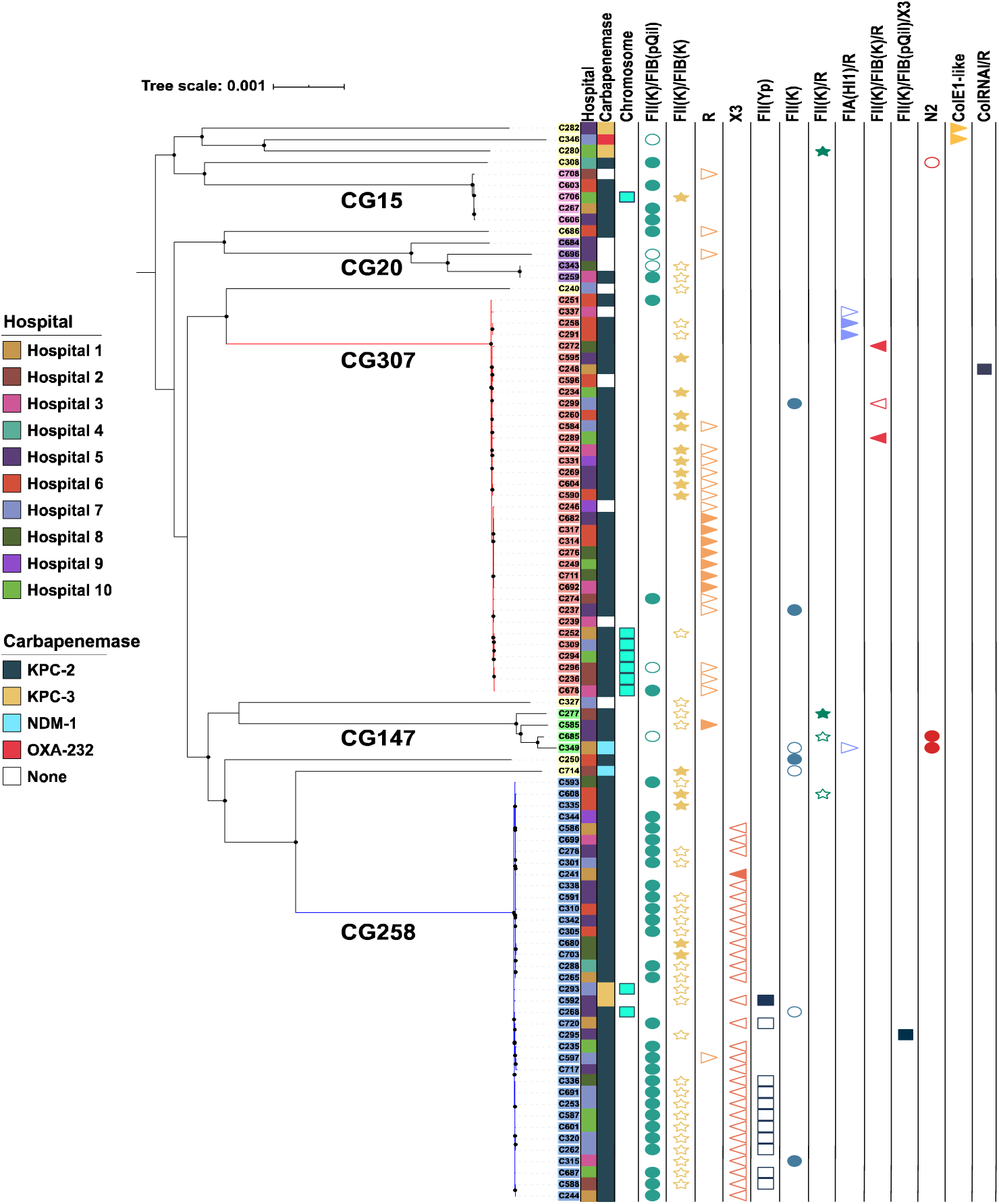
Core Gene Pangenomic Population Structure of 94 CRKP isolates in the Houston, TX cohort. Maximum likelihood phylogenetic tree demonstrating the two predominant co-circulating clades, CG258 (blue clade) and CG307 (red clade). Internal node bootstrap values of ≥ 95% are denoted as black circles. Label backgrounds indicate the 6 hierarchical clustering groups identified using Bayesian analysis of population structure. Clonal groups associated with each hierarchical group are as follows: CG258 (blue); CG307 (red); CG147 (green); CG15 (pink); CG20 (purple); other CGs (yellow). First column indicates each of the ten hospital sites in Houston, TX. Second column indicates carbapenemase carriage. The third column indicates if carbapenemase carriage was chromosomal. The final 13 columns indicate the plasmid vectors that were identified to have at least one sample with carbapenemase carriage. Carbapenemase carriage of each plasmid vector is distinguished by carbapenemase positive (filled shape) and carbapenemase negative (empty shape).

Molecular epidemiological features of the Houston *K. pneumoniae* isolates are shown in Table 1. The median genome size of isolates belonging to CG307 was smaller than CG258 and other heterogeneous sequence types, although there were no significant differences in chromosome sizes between the three CGs. The smaller genome size of CG307 was due to a smaller number of plasmids as compared to CG258 or other CGs (Table 1). The mean number of acquired antimicrobial resistance (AMR) genes per genome, *i.e.* the average number of non-intrinsic AMR genes found per CR*Kp* isolate, was 7.8 ±5.0 genes (Additional file 2: Fig. 2A). The median number of acquired AMR genes stratified by group was similar between CG258 (5; IQR = 6), CG307 (7; IQR=5), and other CGs (9; IQR=10.5). We did not find a statistically significant difference across these subsets in the numbers of genes encoding AMR determinants of different classes (*i.e.,* number of antibiotic classes with at least one resistance determinant per genome) (Additional file 2: Fig. 2B). We also analyzed AMR and virulence determinants using the Kleborate composite resistance scoring metric (30) along with the categorical virulence score (see materials and methods). We found no statistically significant difference in Kleborate resistance scores between CG258 and CG307 (adj. p-value = 0.3). However, CG258 isolates had a statistically significant higher resistance score than the group of other heterogenous sequence types (adjusted p-value = 0.007). The Kleborate composite virulence scores in Table 1 reflect that the Houston CG307 isolates lacked genes encoding siderophores (e.g., yersiniabactin, salmochelin, aerobactin), the genotoxin colibactin gene cluster and hypermucoidity genes (*rmpA* and *rmpA2*), which are commonly found virulence determinants in hypervirulent lineages of *K. pneumoniae sensu stricto* (28, 29).

**Fig. 2.**
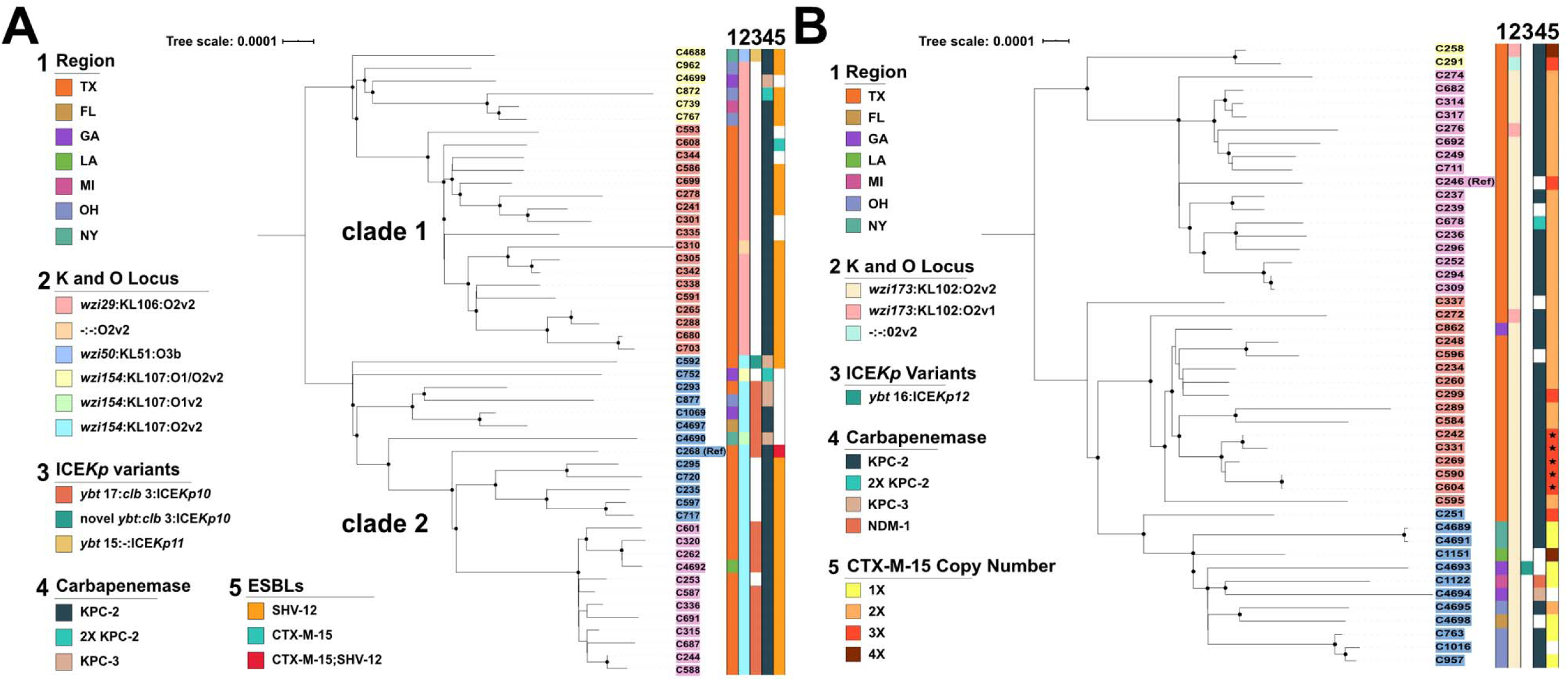
Core SNP inferred ML phylogenies of CG258 and CG307 isolates. Internal node bootstrap values of ≥ 95% are denoted as black circles. Isolate label backgrounds indicate hierarchical clustering groups identified using Bayesian analysis of population structure for each respective sub-lineage. Legend designations are as follows: (1) region from which isolate was collected, (2) capsular synthesis/LPS allele type, (3) ICE*Kp* variant with yersiniabactin and colibactin gene cluster lineages listed respectively, (4) carbapenemase carriage status, (5) ESBL carriage (CG258)/*bla*_CTX-M-15_ copy number (CG307). **(A)** CG258 core SNP phylogeny using a reference-based alignment with C268 isolate. **(B)** CG307 core SNP phylogeny using reference-based alignment with C246 isolate. Stars within *bla*_CTX-M-15_ copy number column (*i.e.* column 5) indicate isolate with one truncated *bla*_CTX-M-15_ copy.

**Table 1.** Pangenomic Features of K. pneumomiae

| **Feature** | **CG258**<br>**(n = 37)** | **CG307**<br>**(n = 35)** | **Other CG**<br>**(n = 23)** | **p-value<sup>a</sup>** |
| --- | --- | --- | --- | --- |
| **genome size (IQR)** | 5,695,223<br>(321,076) | 5,551,257<br>(142,518) | 5,666,705<br>(119,114) | 0.0001 |
| **chromosome size (IQR)** | 5,322,183<br>(187,234) | 5,357,994<br>(15,382) | 5,302,444<br>(155,756) | 0.3 |
| **GC content (IQR)** | 57.1 (0.2) | 57.3 (0.1) | 57.2 (0.2) | <0.0001 |
| **coding genes (IQR)** | 5,380 (269) | 5,158 (139) | 5,286 (120) | <0.0001 |
| **plasmids (IQR)** | 4 (2) | 2 (1) | 4 (3) | <0.0001 |
| **resistance genes (IQR)** | 5 (6) | 7 (5) | 9 (11) | 0.3 |
| **resistance classes (IQR)** | 6 (3) | 6 (1) | 6 (7) | 0.3 |
| **resistance score (SD)<sup>b</sup>** | 2.1 (0.2) | 1.9 (0.4) | 1.6 (0.8) | 0.01 |
| **virulence score (SD)<sup>b</sup>** | 0.7 (1.0) | 0 (0.0) | 0.7 (1.4) | 0.0006 |

CG – clonal group ; IQR – interquartile range; SD – standard deviation; median values are reported with IQR; means are reported with SD
<sup>a</sup>Kruskal-Wallis rank sum test p-value
<sup>b</sup>Composite antimicrobial resistance and virulence scores based on scoring schema developed by Holt et al. Kleborate tool (https://github.com/katholt/Kleborate)

Multiple conjugative elements (ICE*Kp*) carrying virulence factors integrated in the chromosome of *K. pneumoniae* lineages have been previously described (28). These mobile genetic elements largely contribute to hypervirulent, high-risk strains of *K. pneumoniae* commonly observed in Asian countries (5). Interestingly, 35.1% (13/37) of CG258 Houston, TX isolates carried a chromosomally inserted ICE*Kp* with an associated yersiniabactin gene (*ybt*) cluster. The ICE*Kp* in most of these CG258 isolates harbored an ICE*Kp*10 – *ybt*17 sequence type with the exception of one Houston CG258 isolate (C592) that carried a novel *ybt* sequence type. All Houston CG258 isolates harboring ICE*Kp* also contained the ICE*Kp*-associated genotoxin and colibactin gene cluster from the *clb*3 lineage. In contrast, all Houston CG307 isolates lacked ICE*Kp* integration. Only one CG307 isolate in the non-Houston isolates (C4693 recovered from a patient in Georgia) harbored a chromosomal ICE*Kp*, which belongs to the ICE*Kp*12 lineage and encodes *ybt*16 sequence type. An important epidemiological observation relating to the population structure of *K. pneumoniae* is the rare convergence of multidrug-resistant (MDR) high-risk lineages with hypervirulent clones (5, 16). We identified two CR*Kp* isolates, C308 (ST23) and C346 (ST231), that harbored aerobactin-encoding *iuc* genes along with *bla*_KPC-2_ and *bla*_OXA-232_, suggesting that the convergence of MDR and potentially hypervirulent clones is occurring in our study population.

### The population structure in co-circulating CG258 and CG307 indicates nested subgroups within each lineage

The pangenome of the full cohort (n=122), using 94 CR*Kp* Houston isolates with 12 CG307 and 12 CG258 isolates collected from other CRACKLE-2 US hospital sites (n=48) plus three references (see material and methods), consisted of 13,049 genes, of which 3,908 (29.9%) made up the core genome, defined as gene groups included in ≥ 99% of the isolate cohort. The overall median nucleotide divergence, a measure of genetic variation within a population based on normalized polymorphism counts, was 0.59%, suggesting that less than 1% of the core genome nucleotide sites were variant sites. This nucleotide diversity is comparable to previously shown measures of genetic variation within the *K. pneumoniae sensu stricto* phylogroup based on core gene alignment (16). Bayesian hierarchical modeling was used to group individual taxa into clusters based on core gene alignment. The population split into 6 hierarchical levels corresponding to five multidrug-resistant clonal groups (CG15, CG20, CG147, CG258, CG307) and a mixture of other heterogeneous sequence types (Fig. 1).

To dissect lineage specific population structures, recombination free, reference-based core SNP maximum-likelihood phylogenetic trees were created for both CG258 and CG307 (Fig. 2). The inter-group median nucleotide divergence of CG258 and CG307 was 0.6%, which was comparable to overall divergence. The intra-group median nucleotide divergence calculated for the CG258 group was 0.013%, a core genome nucleotide diversity expected for a clonal group showing a more homogenous core genome at the clonal group level, compared to the overall species level. We identified two clades within the CG258 lineage (Fig. 2A), split largely by the capsule synthesis locus (*cps*), as previously described (7, 56). Clade 1 (*wzi 29*/KL106) and clade 2 (*wzi 154*/KL107) encompassed 45% and 51% of CG258 *K. pneumoniae* isolates, respectively. The remaining 4% of CG258 isolates had unique capsular synthesis loci and/or a large IS*Kpn26* mediated deletion in this region (a region known to have a high rate of recombination). There were nested population structures within each clade that segregated by geographical site (Houston vs. other United States CRACKLE-2 sites), with a clearer delineation within clade 1. There was a strong association of isolates harboring ICE*Kp10* (primarily the *ybt*17 lineage) with clade 2 isolates (18/25; 72%) not observed in clade 1 isolates (p-value = 5e-6).

The CG307 group (Fig. 2B) was less divergent compared to CG258, with a median nucleotide divergence of 0.004%. There was a marked geographical split of the CG307 group, correlating with the predicted four hierarchical clusters within the core population structure. A majority of CG307 isolates had the same K and O loci (*wzi173* allele associated with the KL102 locus and the O2v1 [3/47] or O2v2 [33/47] loci, respectively), with the exception of one isolate (C291) which had a 28,813-bp IS*Kpn26* associated deletion in the *cps* locus.

To determine the overall population structure of CG307 isolates in the Houston region, relative to historical isolates detected globally, a reference-based alignment maximum likelihood inferred phylogeny was generated with CG307 isolates (n=798), using C234 (the first CG307 isolate in the Houston population), as a reference (Fig. 3). Four hierarchical population structures were detected, with a prominent distinction between three Houston-based ST307 clades (clade I, III, IV) and the worldwide disseminated CG307 clade (clade II) (Fig. 3). Houston-based Clade III isolates shared a common ancestor with Clade IV isolates not shared with the distinct Clade I and Clade II isolates. Thus, phylogenetic reconstruction suggests that three particular lineages of CG307 (clades I, III, and IV) distinct from CG307 found in other parts of the world (Clade II), are currently circulating in Houston hospitals.

**Fig. 3.**
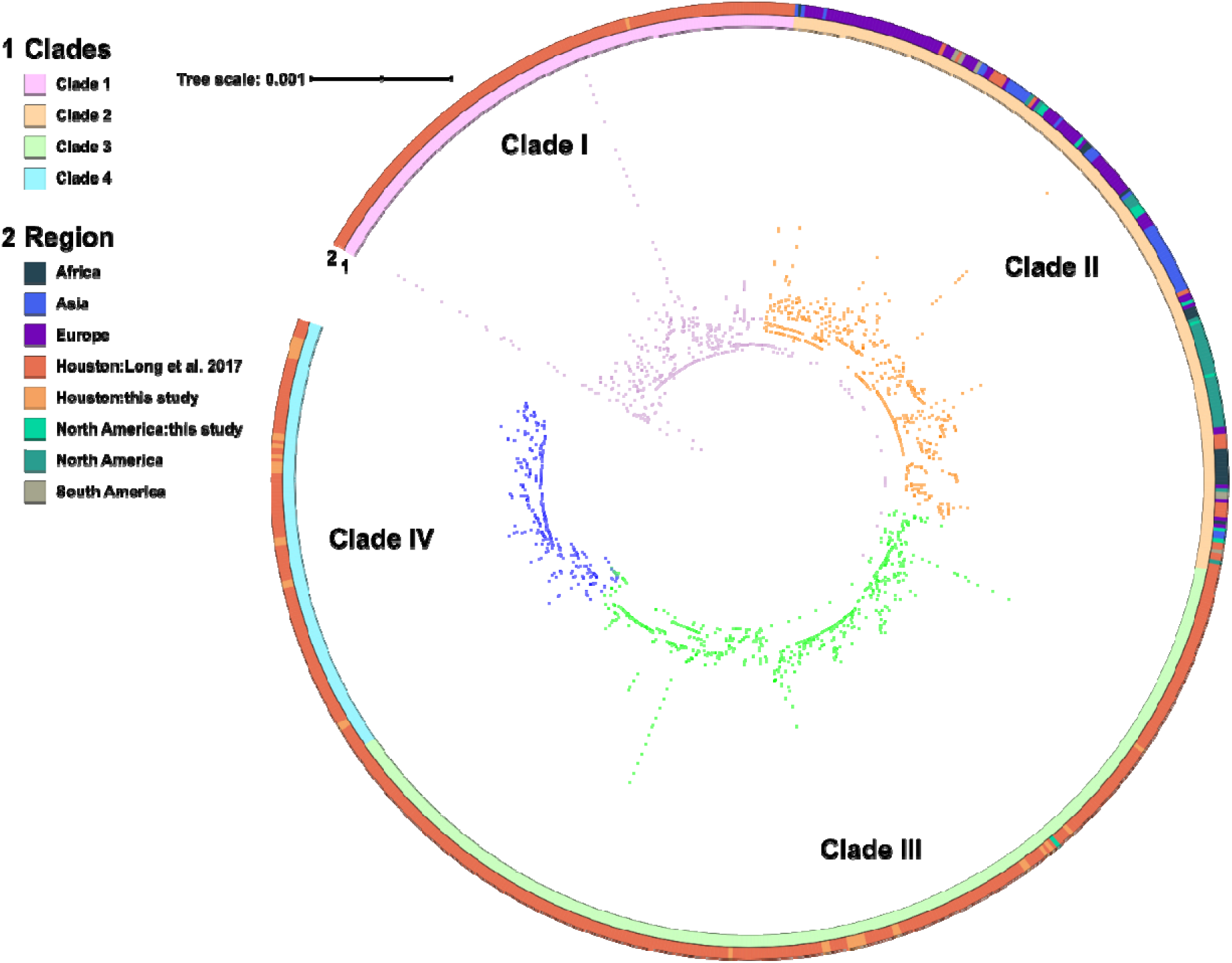
Population structure of previously characterized CG307 isolates with the CG307 Houston CRACKLE II isolates. Maximum likelihood inferred phylogeny of CG307 isolates (n=798) using C234 as a reference for core gene alignment. Branch label background corresponds to hierBAPS predicted clade. Outer ring indicates region where isolate was collected. Clades I, III, and IV are predominantly made up of Houston isolates. Clade II is disseminated worldwide and shares a paraphyletic relationship with Clade I.

### Dissection of the accessory genome suggests independent evolution and limited horizontal gene transfer between CG307 and CG258 lineages

We sought to determine the extent of accessory genome sharing within our CR*Kp* group as a measure of potential interclade horizontal gene transfer of antimicrobial resistance and virulence determinants. We took this approach to understand dynamics of circulation of these genes between high-risk clones that might explain how they co-circulate in the Houston area. Thus, using the full cohort described in the previous section, we sought to determine the geographical clustering of the accessory genomes of all *K. pneumoniae* isolates, including genes encoding carbapenemases. Interestingly, the clustering analysis indicated a distinct separation of the CG258 accessory genome from the rest of the isolates (Fig. 4A and 4B), suggesting that CG258 does not share its accessory genome with other lineages. Fig. 4A shows that, with the exception of one Georgia isolate (C4688), CG307 isolates from Houston cluster together and are distinct from non-Houston CG307. In contrast to the unique accessory genome components in CG307 that segregated by region, we were unable to identify geographical clustering of accessory genome of CG258 isolates.

**Fig. 4.**
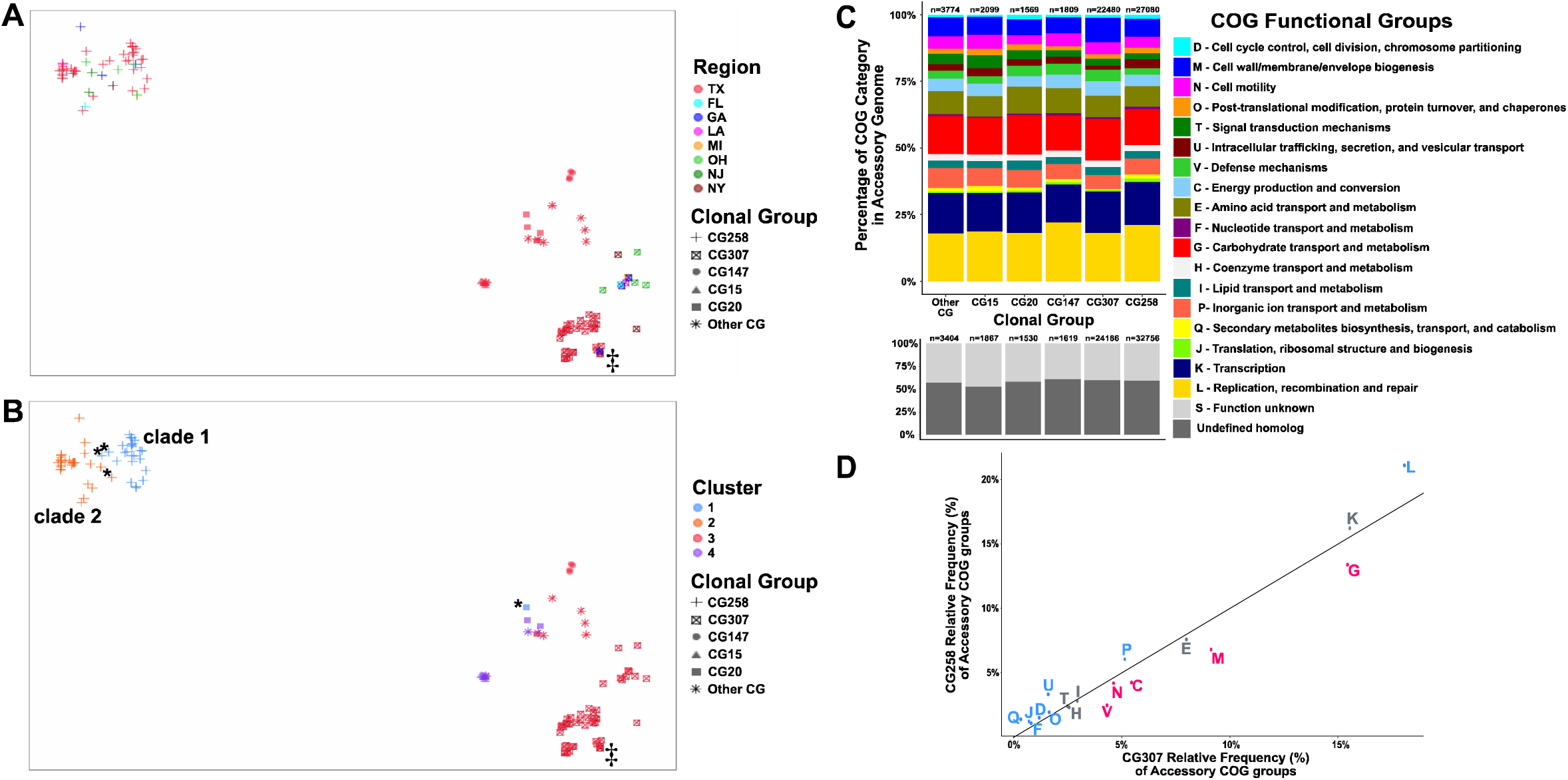
Clustering of CRKP isolates with predicted protein functional characterization of the accessory genome. (**A – B)** t-distributed Stochastic Neighbor Embedding (t-SNE) 2-D plot of accessory genome clustering of CR*Kp* isolates. Clonal group is indicated by shape. ‘‡’ Georgia isolate (C682) that clusters with Houston CG307 isolates. **(A)** geographical region stratification of isolates. **(B)** Cluster group prediction (k=4) using a PAM algorithm to determine cluster assignment. ‘*’ Exceptions to CG258 cluster-to-clade correlation. **(C)** Stacked bar-chart of Cluster of Orthologous Genes (COGs) functional category proportions based on annotated genes found in the accessory genome. ‘n’ above each group indicates number of genes identified in each respective clonal group. Proportion of functionally annotated accessory genes differ significantly across pairwise comparisons of clonal groups as demonstrated in scatter plot **(D)** which plots CG258 vs CG307 relative frequency proportions of COG functional groups denoted by the COG group letter (defined in C) with significant adjusted p-values indicated for greater proportion of CG258 (blue) or CG307 (red) for each respective group labelled accordingly. These COG function group proportions exclude ‘S – unknown function’ and ‘undefined homologs’. Non-significant COG functional group differences are labelled in grey.

We then identified the minimized genomic distance between each isolate to determine which isolates were more likely to share similar accessory genome content based on their predicted cluster assignment. Fig. 4B shows that when cluster groups were identified by sequence similarity (pairwise binary distances between the 122 isolates), there was a split in CG258 that largely segregated by *cps-*associated clades (cluster 1 vs cluster 2), with four exceptions (C293, C295, C259, and C4688). CG307 isolates shared a cluster assignment (cluster 3, Fig. 4B) with ten other isolates belonging to non-CG307. Furthermore, CG307 accessory genomes appeared to cluster with CG147 isolates in our cohort. Principal component analysis (PCA) of the accessory genome indicated that 90.7% of the variance in the dataset could be explained in the first two components of the accessory genome with noted separation of CG258 from CG307, as well as the other clonal groups (Additional file 3: Fig. S3) observed in two-dimensional analysis, further supporting our previous clustering results. Overall, our data suggest that the accessory genomes of CG258 and CG307 diverged with a clear separation between CG258 and other clonal lineages.

To further support the divergence of CG258 from CG307 and lack of genomic sharing, we analyzed a subset of the accessory genome excluding low (≤ 5%) and high (≥ 95%) frequency gene groups that are less indicative of horizontal gene transfer within the full cohort and determined distribution differences by cluster of orthologous genes (COGs) functional categories (Additional file 1: Table S7). Fig. 4C shows the overall distribution of accessory genome content annotated by functional group across all clonal groups found. When focusing on COG functional group proportions of CG258 vs CG307 isolates, we found statistically significant differences in relative frequency proportions of each respective COG functional group with all but 5 of the 18 characterized COG functional groups (Fig. 4D). In contrast, there were fewer statistically significant differences observed in the proportion of COG functional groups within the accessory genomes for both CG258 and CG307 compared to each respective other clonal group found in the cohort (Additional file 2: Fig. S4). These findings support the notion that there are accessory genome functional differences between CG307 and CG258 and there have been independent adaptive responses to potential selective pressures.

The largest statistically significant differences in COG functional groups between CG258 and CG307 were in predicted carbohydrate metabolism and transport mechanisms (G), cell membrane structure/biosynthesis (M), and replication/recombination/repair (L) genes (Fig. 3D). The larger proportion of replication, recombination, and repair genes (L) in CG258 is likely due to a higher number of plasmids observed in CG258 compared to CG307 isolates. Noted differences include a previously described chromosomal, 13 kb π conserved in all CG307 isolates (n = 48) and absent from all CG258 (n = 51) (adj. p-value = 8e- 33) (Additional file 2: Fig. S5). The aforementioned fimbrial gene cluster was also present in all four CG147 isolates. A second, previously described (12), chromosomal capsular synthesis cluster with 12 genes (Cp2) was present in all 48 CG307 isolates and absent from all others (adj. p-value = 9e-34) (Additional file 2: Fig. S5). Conversely, there was a carbohydrate metabolism operon (designated *ydjEFGHIJ*) exclusively found on all CG258 and CG147 isolates, and absent in the CG307 isolates (adjusted. p= 5e-34). Carriage of unique phages between CG307 and CG258 was a primary driver of accessory genome differences. Indeed, three intact phages detected in CG307 were absent in CG258 (Additional file 2: Fig. S5). These results collectively indicate that CG258 and CG307 each contain highly conserved and distinct accessory genomes, which are likely driving independent endemic spread of each lineage in the Houston region.

### Carriage of chromosomal *bla*_CTX-M-15_ is the major antimicrobial resistance determinant of CG307 in the Houston area with evidence of independent acquisitions of *bla_KPC-2_*

Table 2 provides an overview of the antimicrobial resistance profile of the CR*Kp* Houston isolates. The number of isolates carrying a carbapenemase gene was 84/95 (88.4%). The majority of the isolates harboring carbapenemase genes were *bla*_KPC-2_ carriers (77/95; 81.1%), followed by *bla*_KPC-3_ (4/95; 4.2%), *bla*_NDM-1_ (2/95; 2.1%) and *bla*_OXA-232_ (1/95; 1.1%). Most *bla*_KPC_ carriage occurred on the Tn*4401a* isoform (94%; Additional file 1: Table S8). CG258 isolates harbored either *bla*_KPC-2_ (35/37; 94.6%) or *bla*_KPC-3_ (2/37; 5.41 %) with the majority on pKpQIL plasmids (27/37; 73%), whereas most CG307 isolates (31/35; 88.6%) carried *bla*_KPC-2_ on different vectors, with one isolate (C678) carrying two *bla*_KPC-2_ copies, one on the chromosome and the other on a FIIK/FIB(pQIL) plasmid. The distribution of *bla*_KPC_-containing plasmids was more diverse in CG307, with evidence of multiple recombination events in plasmids carried by CG307 vs CG258 isolates (Fig. 1), in particular with different F-type and R-type plasmids. Interestingly, we could identify a cluster of six CG307 isolates (average 40 SNPs differences) that harbored Tn*4401a* in the same chromosomal context (Fig. 1) originating from five different Houston regional hospitals.

**Table 2.** Antimicrobial Resistance Features of 95 K. pneumoniae from Houston, TX

| Feature | CG258<br>(n = 37)<sup>a</sup> | CG307<br>(n = 35)<sup>a</sup> | Other CG<br>(n = 23)<sup>a</sup> | p-value<sup>b</sup> |
| --- | --- | --- | --- | --- |
| **Carbapenemases** |  |  |  | <0.0001 |
| *bla*<sub>KPC-2</sub> | 35 (94.6%) | 31 (88.6%) | 11 (47.8%) |  |
| *bla*<sub>KPC-3</sub> | 2 (5.4%) | 0 (0.0%) | 2 (4.2%) |  |
| *bla*<sub>NDM-1</sub> | 0 (0.0%) | 0 (0.0%) | 2 (4.2%) |  |
| *bla*<sub>OXA-232</sub> | 0 (0.0%) | 0 (0.0%) | 1 (1.1%) |  |
| **ESBLs** |  |  |  | <0.0001 |
| *bla*<sub>CTX-M-15</sub> | 1 (2.7%) | 35 (100%) | 11 (47.8%) |  |
| *bla*<sub>SHV-12</sub> | 30 (81.1%) | 0 (0.0%) | 0 (0.0%) |  |
| *bla*<sub>SHV-27</sub> | 0 (0.0%) | 0 (0.0%) | 1 (4.3%) |  |
| *bla*<sub>SHV-12</sub>/*bla*<sub>CTX-M-15</sub> | 1 (2.7%) | 0 (0.0%) | 0 (0.0%) |  |
| *ompK35/ompK36* changes<sup>c</sup> | 37 (100%) | 35 (100%) | 5 (21.7%) | <0.0001 |
| **Aminoglycoside<sup>d</sup>** | 36 (97.3%) | 30 (85.7%) | 15 (65.2%) | 0.003 |
| **Tetracycline<sup>d</sup>** | 2 (5.4%) | 3 (8.6%) | 9 (39.1%) | 0.002 |
| **Sulfonamide<sup>d</sup>** | 19 (51.4%) | 16 (45.7%) | 14 (60.9%) | 0.5 |
| **Phenicol<sup>d</sup>** | 14 (37.8%) | 8 (22.9%) | 9 (39.1%) | 0.3 |
| **Fluoroquinolone<sup>e</sup>** | 37 (100%) | 35 (100%) | 14 (60.9%) | <0.0001 |
| **Colistin<sup>e</sup>** | 2 (5.4%) | 3 (8.6%) | 1 (4.3%) | 0.9 |

<sup>a</sup>Categorical responses include counts and percentages in parenthesis.
<sup>b</sup>Fisher's exact test p-value.
<sup>c</sup>Outer membrane porin gene changes are counted if a truncation or nonsense mutation is detected in either the *ompK35* or *ompK36* gene.
<sup>d</sup>Aminoglycoside, tetracycline, sulfonamide, and phenicol predicted resistance based on presence of at least one gene that confers resistance to respective antibiotic class (e.g. *aac3-Ia* confers aminoglycoside resistance).
<sup>e</sup>Fluoroquinolone and colistin predicted resistance based on chromosomal mutations that confer resistance to each respective antibiotic class (e.g. truncations in *mgrB/pmrB* confer resistance to colistin). Small proportion of isolates (6/95; 6.3%) also carry quinolone resistance genes (i.e. *qnr* variants).

A characteristic feature of the Houston CG307 is that all isolates harbored more than one copy of *bla*_CTX-M-15_ (2 [n=25], 3 [n=9] and 4 [n=1]) (Table 2 and Fig. 2B). *bla*_CTX-M-15_ was primarily located in the chromosome, with 2-3 copies per genome in relatively similar genomic contexts. Only 3 Houston CG307 isolates had plasmids (pC251_1, pC258_1, pC291_1) harboring *bla*_CTX-M-15_. All 3 plasmids were F-type pKPN3-like plasmids, which have been associated with CG307 isolates in other geographical locales (10, 11). In contrast to CG307, only 2/37 (5.4%) CG258 isolates harbored *bla*_CTX-M-15_, with the two isolates having the gene on distinct pKPN3-like plasmids (pC268_1 and pC608_2). The *bla*_SHV-12_ variant was the most common ESBL gene carried by CG258.

Except from a CG307 Georgia isolate (C862) which shared similar genomic characteristics with CG307 from Houston, all other CRACKLE-2 CG307 recovered outside of Houston exhibited distinct accessory genome features (Fig. 2B), supported by previous phylogenetic analyses (see above). Indeed, a majority of non-Houston CG307 (10/12, 83%) harbored a pKPN3-like plasmid with 6 having *bla*_CTX-M-15_ as part of an IS*Ecp1* element. Furthermore, 3 of the 8 CG307 non-Houston isolates carried *bla*_KPC_ on non-Tn*4401a* transposons (compared to all Houston isolates, which carried *bla*_KPC_ on Tn*4401a*; Additional file: Table S8). Additionally, non-Houston CG307s harbored A/C-type plasmids (n=2) and carrying *bla*_KPC_, a feature absent in Houston isolates (Fig. 5). These results support that a unique and distinct epidemic lineage of CG307 is currently circulating in tertiary care hospitals in Houston, TX.

**Fig. 5.**
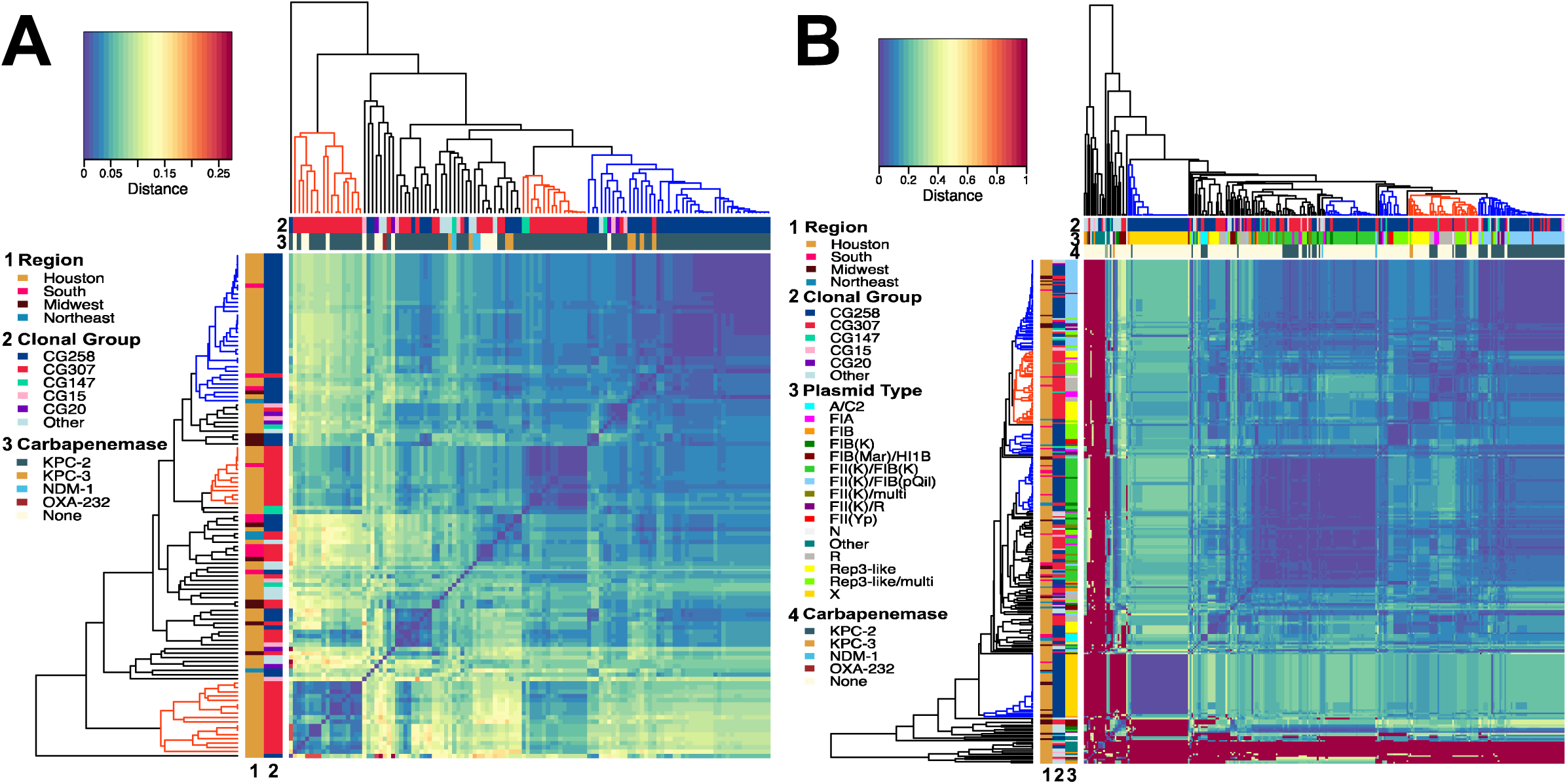
Dendrogram and Heatmap of Plasmidome Mash distances. Legend for row and column labels listed to the left of each respective graph. Each dendrogram is constructed through agglomerative hierarchical clustering using an ‘average’ linkage. **(A)** Plasmidome mash distance matrix by isolate (n = 119). Legend is labeled as follows: (1) Region; (2) Clonal Group; (3) Carbapenemase. There is a noted primary clustering group of CG258 isolates (blue labeled clade) whereas there are two Houston-based CG307 clustering groups (red labeled clades) with diffuse clustering occurring with other CGs. **(B)** Plasmidome mash distance matrix by plasmid type (n = 295) with small, primarily ColE1-like plasmids excluded from analysis. Legend is labeled as follows: (1) Region; (2) Clonal Group; (3) Plasmid Type; (4) Carbapenemase. This analysis shows clustering of primarily CG258 X2 type plasmids, multi-replicon FIIK type plasmids, and pKpQIL plasmids indicated with blue labelled cluster groups. This is contrast to one primary CG307 plasmid cluster group which includes R type plasmids as well the novel pCG307_HTX plasmid associated with the Houston group.

### CG258 and CG307 isolates harbor unique plasmid content

To determine the full complement of plasmid vectors assembled in our CR*Kp* cohort, we characterized the full plasmid content (i.e., the plasmidome) to assess potential clustering and sharing of plasmids by clonal group. The plasmidome of our CR*Kp* isolates was highly diverse (Fig. 5) with an average number of plasmid structures of 3.5 per isolate (min 1; max 7). Plasmid content clustered by core-defined clonal groups (Fig. 5A) with a majority of CG258 isolates clustering within their own distinct group (Fig. 5A, top right). In particular, there were three plasmid-types associated with the CG258 lineage, including X3, FII(K)/FIB(K) and FII(K)/FIB(pQIL) plasmids (Fig. 5B). There was a particular rep-3 family replication initiator protein (52/415; 12.5%) associated with a plasmid (rep_cluster_1418; Additional file 1: Table S4) found in nearly all Houston CG307 isolates (31/35; 88.8%). These CG307 rep-3 family plasmids (herein referred to as pCG307_HTX) primarily clustered with F-type and R-type plasmids (Fig. 5B). 30/52 (57.7%) pCG307_HTX plasmids were predicted to be non-mobilizable, with the rest (22/52; 42.3%) predicted to be conjugative and/or mobilizable, harboring *tra* operons and/or *mob* encoding genes with essential *oriT* sites. Importantly, 10/31 (32.3%) pCG307_HTX plasmids carried Tn*4401a*-*bla*_KPC_ (Fig. 5B), suggesting that this plasmid may have been important for carbapenemase dissemination. Collectively, our results indicate that the CG258 plasmidome is conserved and largely segregated from other co-circulating CR*Kp* isolates, including CG307, in agreement with the pan-genome analyses.

### The novel pCG307_HTX plasmid carrying Tn4401a conjugates at similar rates compared to CG258 pKpQIL plasmid

To evaluate the potential for horizontal transfer of carbapenemase genes, we chose 4 plasmid candidates of interest based on plasmidome results, with three belonging to CG307 and one to CG258. We assessed relative conjugation transfer frequencies of two *bla*_KPC-2_ carrying plasmids with the novel rep-3 family *repA* gene (*i.e.,* pCG307_HTX plasmids, one each predicted to be conjugative [pC299_2] or not conjugative [pC711_1], respectively). Conjugation efficiencies were compared using a comparable size pKpQIL plasmid (pC344_1, CG258 vs pC299_2, CG307) that shared a similar *tra* operon and cargo gene region (Additional file 2: Fig. S6). Additionally, we chose a pKPN3-307_typeA-like plasmid (pC763_2) primarily found in our non-Houston CG307 isolates to compare its conjugative frequency with the aforementioned F-type plasmids (Additional file 2: Fig. S6). Three of the four F-type plasmids had detectable, positive transconjugants in a proportion of conjugation transfer assay experiments (Additional file 2: Fig. S7), with varying efficiencies. pKpQIL (pC344_1) and rep-3 family multi-replicon F-type (pC299_2; pCG307_HTX) plasmids had comparable, low average transfer frequencies of 4.1 × 10^-7^ and 2.9 × 10^-6^, respectively. The pKPN_3_typeA plasmid associated F-type plasmid had a greater transfer frequency (6.3 × 10^-5^). However, this did not reach a statistically significant difference (p = 0.06). As expected through *in silico* prediction, pC711_1 yielded no transconjugants in any of the three conjugation transfer assays. Overall, our conjugation experiments validated *in silico* predictions of mobilization and confirmed potential broad host range transmission of the novel pCG307_HTX plasmid associated with the Houston CG307.

### Patients with CG307 colonization or infection may have more favorable outcomes compared to those with CG258

Clinical epidemiological features stratified by CG for the Houston, TX isolates are presented in Table 3. The crude 30-day mortality for the full Houston cohort (n=73) was 17.8%; (13/73 [95% CI: 7.8 – 22.6%]). When comparing clinical features across the CG258 and CG307 groups, patients with CG258 exhibited a statistically significant higher 30-day mortality compared with those infected/colonized with CG307, albeit with a small number of events. Patients with CG307 infection/colonization exhibited a higher proportion of samples isolated from urine (65.6%) compared to CG258 (37.0%), but this association did not reach statistical significance (p=0.068). Conversely, patients with CG258 infection/colonization had a higher proportion of isolates from blood (14.8% vs 0%) and respiratory cultures (25.9% vs 12.5%) compared to the CG307 patient group.

**Table 3.**
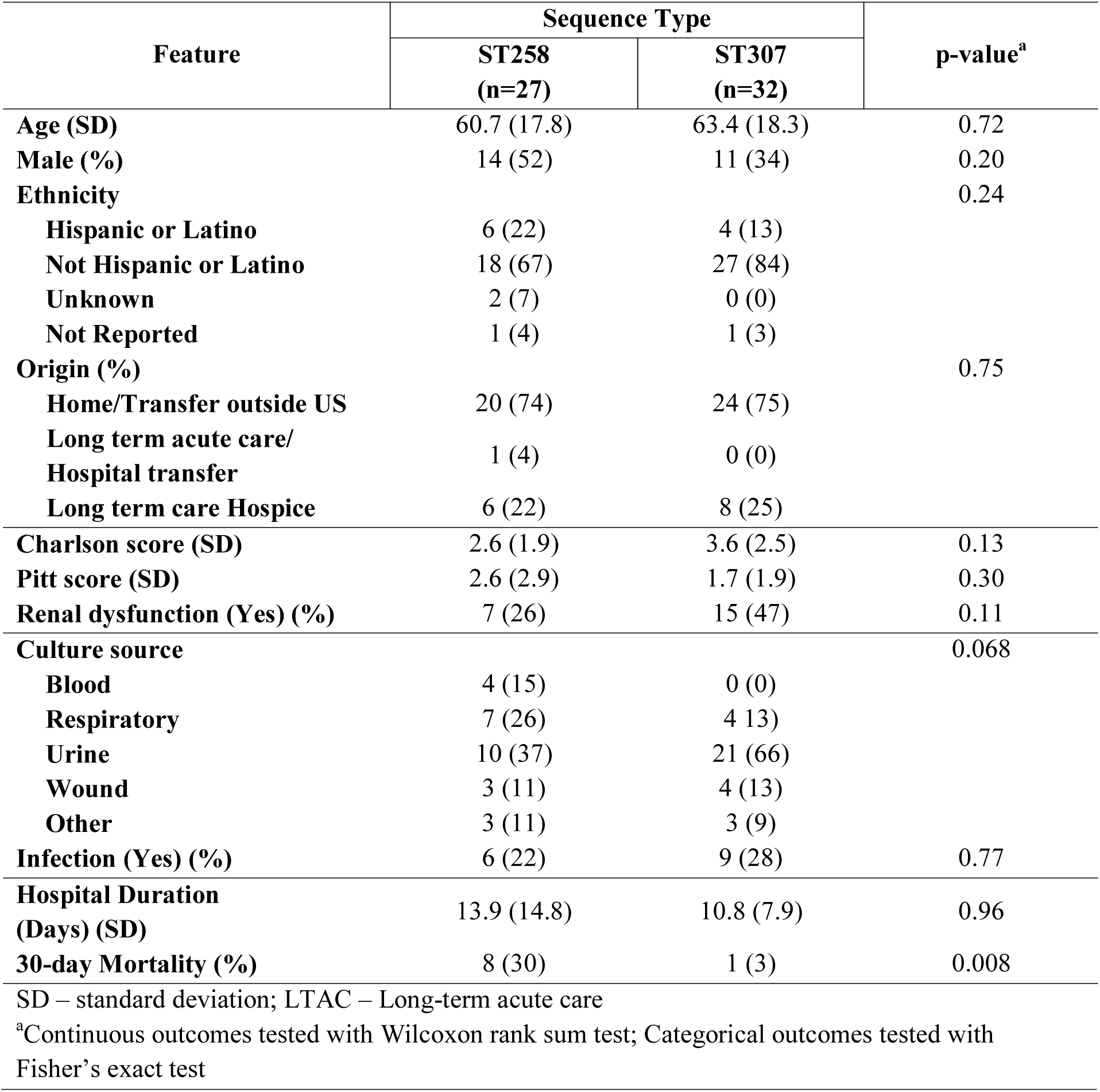
Clinical Features and Outcomes of CG258 vs CG307 from Houston, TX

To further investigate differences in clinical outcomes for patients with CG307 vs CG258, we calculated an inverse probability weighted (IPW) Desirability of Ordinal Outcome Ranking (DOOR) outcome (55) at 30-days (see Materials and Methods) for the Houston, TX cohort. The IPW-adjusted DOOR probability estimate was 64% (95% CI: 48-79%). This represents the probability that a randomly selected CG307 patient would have a better outcome than a randomly selected CG258 patient, and suggests that patients with CG307 colonization or infection trended towards better overall outcomes following adjustment for potential confounding that may arise in an observational study (Table 4).

**Table 4.**
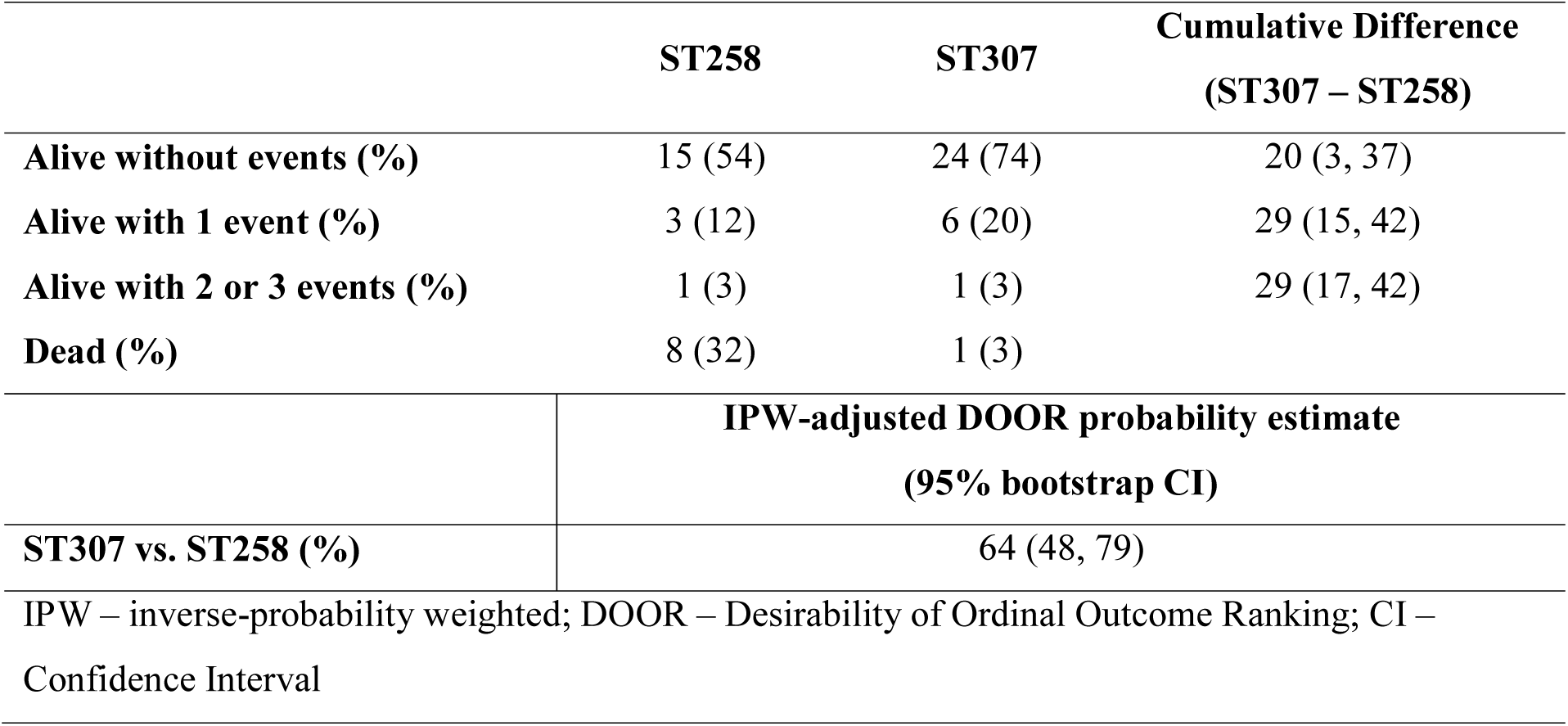
IPW-adjusted DOOR categories for Houston, TX

| Table 4. IPW-adjusted DOOR categories for Houston, TX |  |  |  |
| --- | --- | --- | --- |
|  | ST258 | ST307 | Cumulative Difference<br>(ST307 – ST258) |
| Alive without events (%) | 15 (54) | 24 (74) | 20 (3, 37) |
| Alive with 1 event (%) | 3 (12) | 6 (20) | 29 (15, 42) |
| Alive with 2 or 3 events (%) | 1 (3) | 1 (3) | 29 (17, 42) |
| Dead (%) | 8 (32) | 1 (3) |  |
|  | IPW-adjusted DOOR probability estimate<br>(95% bootstrap CI) |  |  |
| ST307 vs. ST258 (%) | 64 (48, 79) |  |  |
| IPW – inverse-probability weighted; DOOR – Desirability of Ordinal Outcome Ranking; CI – Confidence Interval |  |  |  |

## Discussion

Although *K. pneumoniae* CG258 has been considered the major genetic lineage responsible for endemicity of carbapenem resistance, in recent years, a new lineage, designated *K. pneumoniae* CG307, has emerged (11–13). Since the first report of CG307 in the Netherlands in 2008, it has been identified in different parts of the world, including Colombia (57), Italy (58, 59), South Africa (60, 61), Pakistan (62) and South Korea (63), among others. Interestingly, the first detection of CG307 in the United States occurred in Houston, TX in 2013 (64) and was subsequently followed by an outbreak in a large, Houston hospital system (15). A more recent study assessing the clinical and genomic epidemiology of carbapenem-resistant *Enterobacterales* in the United States (3), showed that Houston was the first major city in the US where carbapenem-resistant *K. pneumoniae* CG307 seemed to have established endemicity along with isolates belonging to CG258. Furthermore, a recent study suggests that CG307 may be spreading to other municipalities in south Texas and co-circulating with CG258 (65). The concomitant circulation of these major clones provided an opportunity to dissect their dynamics of dissemination, genetic relationships, and evolution using a pangenomic approach combined with robust clinical data. Our findings suggest that CG258 tends to have a higher median plasmid content with a similar number of AMR determinants found across all CR*Kp* isolates. The absence of any known determinants associated with hypervirulent lineages in CG307 suggests that CG258 may have a higher likelihood of causing more adverse infections through the convergence of high-risk multidrug resistant and hypervirulent genotypes in our study population. Of note, this assumption was supported by our DOOR analyses.

The most striking finding of our study was that the two main multidrug-resistant *K. pneumoniae* lineages have evolved independently of one another and appear to be disseminating in parallel with limited evidence of inter-clade horizontal gene transfer between them. Moreover, our results support the notion than CG307 plasmids carrying KPC carbapenemases are likely to be shared with other clonal groups, except CG258, amplifying the epidemic of multidrug-resistant organisms in the same geographical area. Partitioning of the accessory genome by COG functional groups indicate large differences in distribution between CG258 and CG307 accessory genome content that are less apparent when comparing each respective clade with other clonal groups. These differences are potentially driven, in part, by CG307 having greater proportion of carbohydrate metabolism and cell membrane synthesis determinants. Unique virulence factors found in our cohort, such as a separate chromosomal capsular synthesis locus (Cp2) shared across CG307 isolates and plasmid-borne glycogen synthesis clusters found on CG307 plasmids have been previously documented in CG307 isolates found in Colombia and Italy (12).

Virulence factors such as the filamentous, extracellular organelles and type 1 fimbriae are associated with colonization and infection of the urinary tract in *E. coli* and *K. pneumoniae* (66–69). Interestingly, a unique π-fimbriae gene cluster (12) was strongly associated with CG307 and CG147 isolates, with urine as a main source where CG307 were recovered. Furthermore, we found accessory genome intersection between CG307 and CG147 isolates by clustering analysis, with comparable proportions of COG functional groups carried by each group. Indeed, there has been recent evidence of parallel antimicrobial resistance patterns shared between CG307 and CG147 with acquisition of similar F- and R-type plasmids harboring *bla*_CTX-M-15_ along with identical *gyrA* and *parC* mutations conferring quinolone resistance, suggesting a common evolutionary pathway (12).

Our study also identified a unique plasmid (designated pCG307_HTX) in CG307 isolates carrying a rep-3 family initiator replication protein with potential for vertical and horizontal transmission based on predicted mobility, as well as positive transconjugants in our conjugative transfer assays. A similar rep-3 family plasmid that recombined with an FII(K) plasmid had been described in a CTX-M-15 associated ST416 *K. pneumoniae* isolate (pKDO1; GenBank accession #: JX424423) (70). We also found that the previously identified FII(K)/FIB(K) plasmid(12), harboring *bla*_CTX-M-15_ associated with worldwide CG307 isolates, had relatively higher conjugative transfer efficiency compared to the Houston pCG307_HTX and CG258 pKpQIL plasmid. This FII(K)/FIB(K) plasmid harboring *bla*_CTX-M-15_ was less prevalent in the Houston CG307 cohort (n=3) where the primary vector of ESBL transmission was stable integration of two copies of a chromosomal IS*Ecp1*-*bla*_CTX-M-15_ transposon. We found that while CG258 may have a greater number of plasmids per genome, plasmidome analyses suggest a greater diversity of unique vectors carrying genes encoding carbapenemases for the CG307 lineage with the potential for sharing across other circulating non-CG258 CR*Kp* isolates in the Houston region.

Our clinical data from patients infected/colonized with carbapenem-resistant CG258 vs CG307 in Houston, TX suggests that patients harboring CG307 may have more favorable outcomes compared to those with CG258 although our DOOR probability estimate didn’t reach statistical significance. Genomic insights provided from our analyses also support CG307 appearing to carry fewer hypervirulence determinants than CG258 isolates. Thus, it is tempting to speculate that the CG307 lineage in our population has evolved as a human colonizer with the propensity to acquire and disseminate antibiotic resistance determinants to other lineages (except CG258), perpetuating and amplifying dissemination of genes coding for carbapenemases. Interestingly, this propensity for disseminating multiple AMR determinants to other clonal groups was noted in a recent outbreak of CG307 in northeast Germany (71). In contrast to our study, they had also found the potential of convergent hypervirulent and multi-drug resistant CG307 isolates due to promiscuous sharing of plasmids carrying multiple hypervirulent and AMR determinants (71). Our epidemiological investigation along with this northeast Germany surveillance study indicate the high potential of horizontal gene transfer along with noted regional variation in accessory genome content across different CG307 clades.

There are limitations in our study. Caution should be exercised when inferring differences in clinical outcomes of patients with CG258 vs. CG307 due to inclusion of both infection and colonizing isolates with small sample sizes. Additionally, there may be underlying characteristics in the two respective patient populations that were not accounted for in our DOOR analysis. While generalizability may only apply to the Houston region, additional sampling will be required to determine if these clinical outcome results persist in other regions where CG307 may be circulating. Nonetheless, despite these limitations, the combination of our IPW-DOOR analysis along with our genomic data suggest that such differences in clinical outcomes may be meaningful in our patient population.

## Conclusions

In conclusion, our data provide a detailed dissection of a unique phenomenon of parallel epidemics of two high-risk lineages of carbapenemase-producing *K. pneumoniae*, both of which are considered an urgent public health. The genomic and clinical insights presented here are likely to provide novel understanding of the epidemiology of multidrug-resistant *K. pneumoniae* in order to design innovative tools to improve patients’ outcomes.

## Data Availability

Short-read and long-read fastq files along with complete assemblies for all K. pneumoniae isolates sequenced including isolates previously published(3) are in BioProject PRJNA658369. The supplemental datasets supporting the conclusions of this article are included within the article and its additional files. Additional files supporting the conclusions of this article are available in a Gitlab repository, 
https://gitlab.com/carmig_dissertations/shropshire_dissertation/crkp_supplemental_files.

https://gitlab.com/carmig_dissertations/shropshire_dissertation/crkp_supplemental_files.

## List of Abbreviations

CG: Clonal Group
COG: Cluster of Orthologous Genes
CR*Kp*: Carbapenem-resistant *Klebsiella pneumoniae*
DOOR: Desirability of Ordinal Outcome Ranking
ESBL: Extended-spectrum Beta-lactamase
IPW: Inverse Probability Weighted
PCA: Principal Component Analysis
ST: Sequence Type
t-SNE: t-Distributed Stochastic Neighbor Embedding

## Declarations

### Ethics Approval and Consent to Participate

Original data was collected with the approval of The University of Texas Health Science Center at Houston IRB, the Committee for Protection of Human Subjects (CPHS; Protocol ID: HSC-MS-16-0334; Ethical approval: 5/16/2016), and no further data were collected from these subjects. CRACKLE-2 data collected outside of Houston was approved through the Duke University Health System Institutional Review Board for Clinical Investigations (DUHS IRB; Protocol ID: Pro00071149; Ethical approval: 6/29/2016) with no clinical data reported in this study. All clinical isolates were stripped of identifying information prior to analysis. All clinical data was deidentified upon receipt.

### Consent for Publication

Not applicable

### Availability of Data and Materials

Short-read and long-read fastq files along with complete assemblies for all *K. pneumoniae* isolates sequenced including isolates previously published (3) are in BioProject PRJNA658369. The supplemental datasets supporting the conclusions of this article are included within the article and its additional files. Additional files supporting the conclusions of this article are available in a Gitlab repository, https://gitlab.com/carmig_dissertations/shropshire_dissertation/crkp_supplemental_files.

### Author contributions

W.C.S and A.Q.D.: next generation sequencing (NGS). W.C.S.: comparative genomics analysis and bioinformatics. W.C.S., D.P., and S. I. G.-V.: plasmid experiments and analysis. C.H., D.V.D., R.B., and B.K. provided and curated clinical data. W.C.S., L.K., M.E., and H.M.: statistical analyses. W.C.S.: original draft preparation. C.A.A., B.M.H., and S.A.S.: Conceptualization, supervision, and original draft preparation. C.A.A.: procured funding for project. All authors contributed to interpretation of results and critical review of manuscript.

## Acknowledgements

Not applicable

