## Additional file 2 for "Accessory Genomes Drive Independent Spread of Carbapenem-Resistant *Klebsiella pneumoniae* Clonal Groups 258 and 307"

**Supplemental figures**

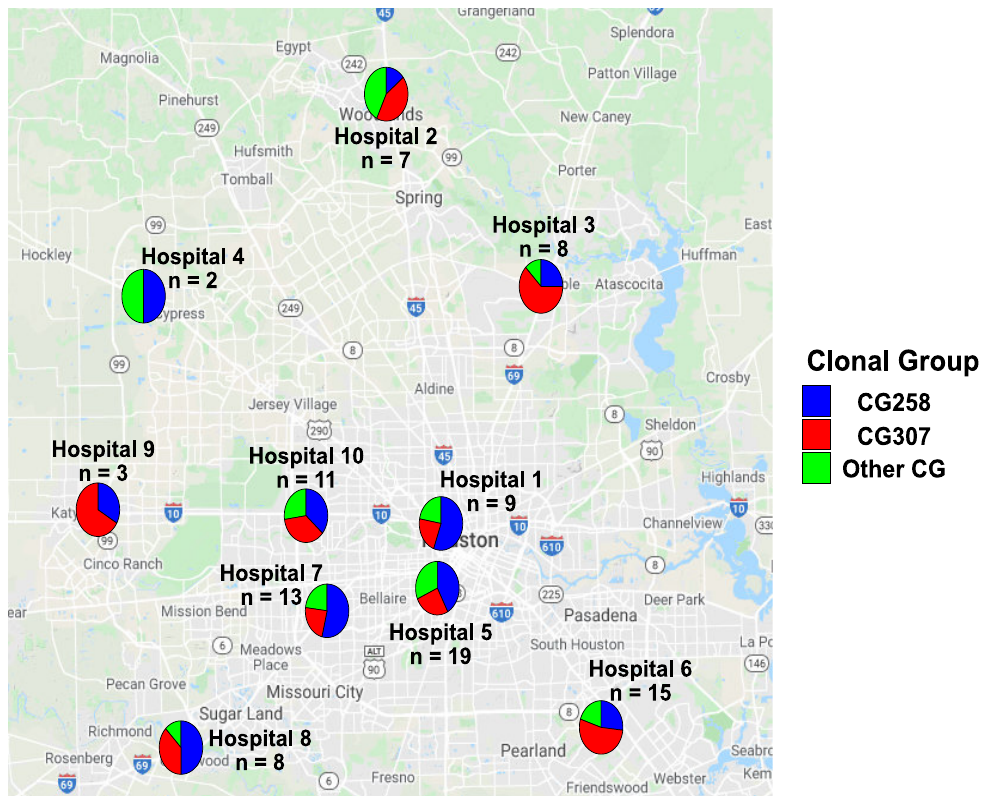
 **Fig. S1.** Geographical distribution of CRKP isolates by clonal groups within a large, Houston, TX region hospital system. There is a comparable distribution of each of the clonal groups across each of the 10 hospital sites in the Houston metropolitan region (Fisher’s exact test: p = 0.8).

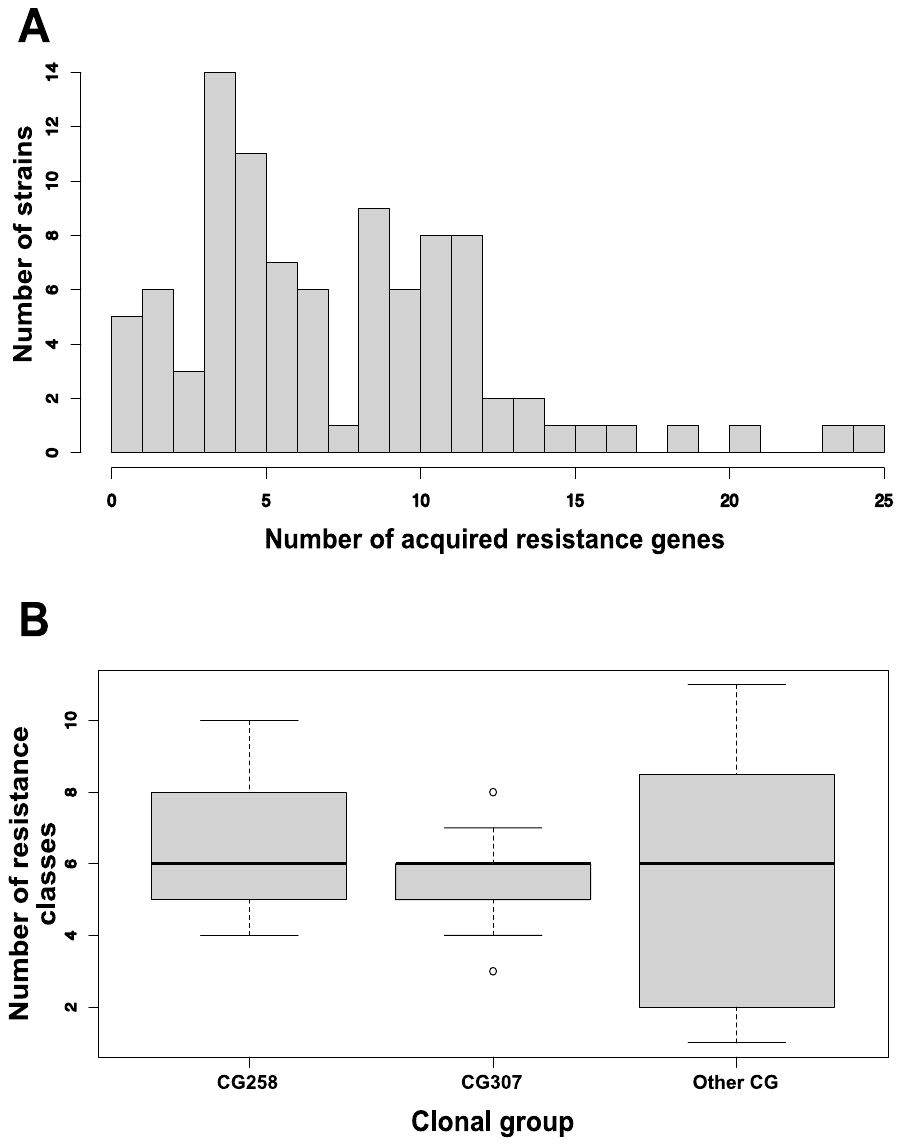

**Fig. S2.** Overall antimicrobial resistance gene acquisition and resistance class characterization. (**A)** Histogram of the number of antimicrobial resistance genes found per isolate genome. **(B)** Box plot of antimicrobial resistance classes identified stratified by sequence type group.

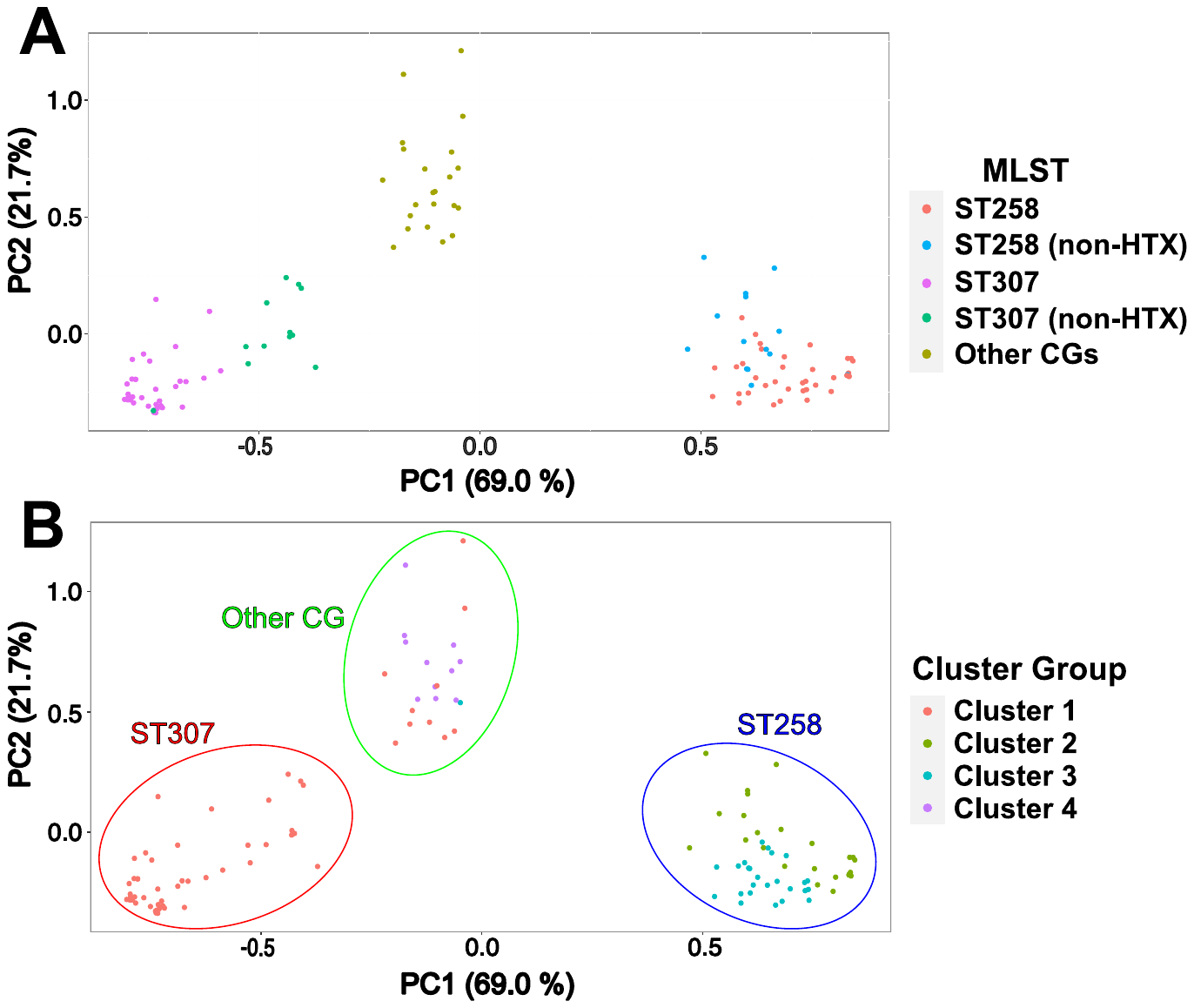

**Fig. S3.** Principal component 2D plot of PC1 and PC2 from the accessory genome gene presence/absence matrix. **(A)** MLST and Houston vs. non-Houston origin; **(B)** Cluster grouping (k=4) as predicting using a PAM algorithm to determine cluster assignment.

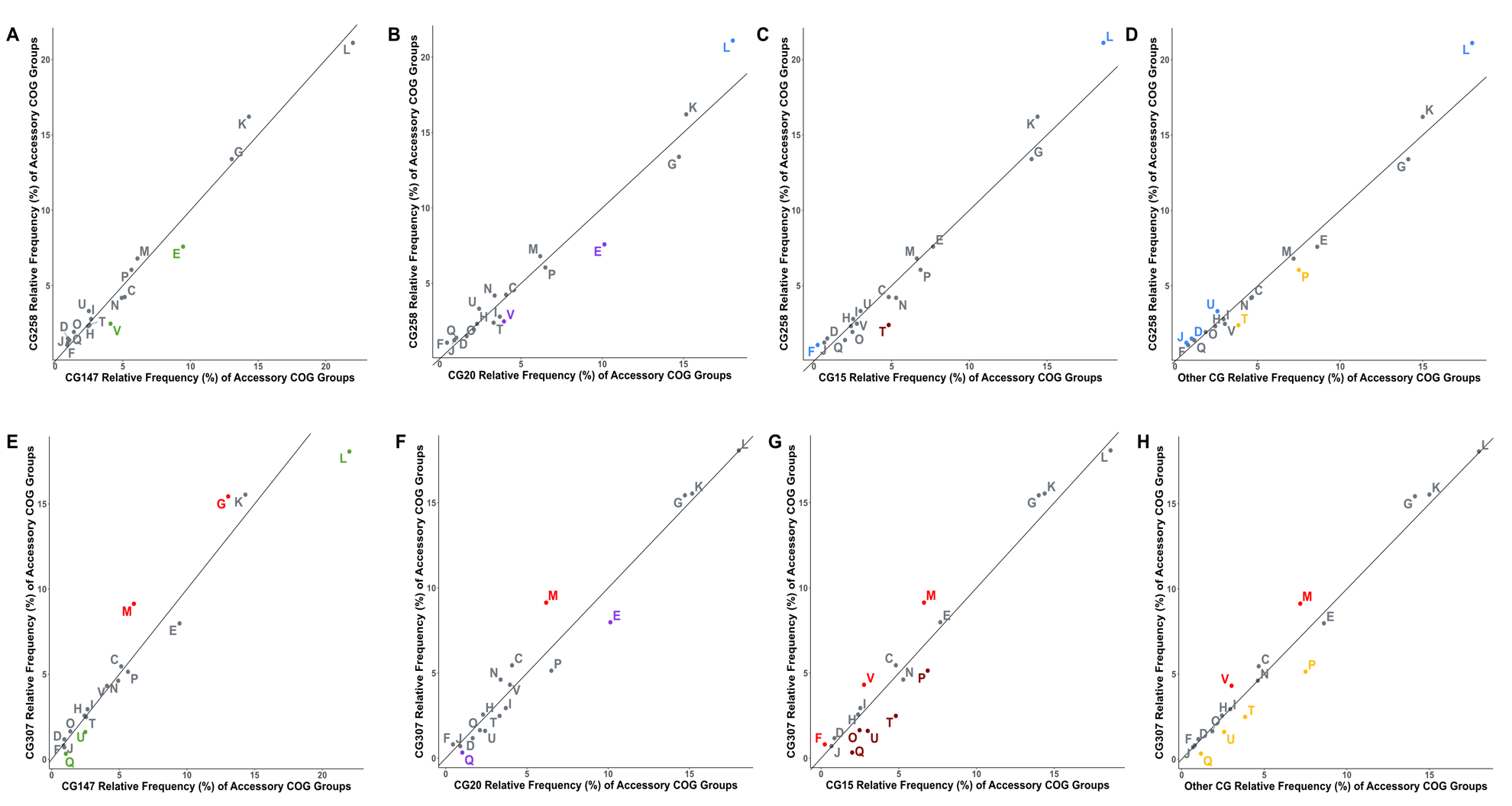

**Fig. S4.** Cluster of Orthologous Gene Functional Group Comparisons of CG258 and CG307 with other clonal groups circulating in Houston, TX. Significant adjusted p-values indicated for greater proportion of CG258 (blue) or CG307 (red) for each respective group labelled accordingly compared to CG147 (green), CG20 (purple), CG15 (pink/dark red), and other CG (yellow) respectively. Non-significant differences are labelled in grey. The proportion excludes ‘S – unknown function’ and ‘undefined homologs’ in the denominator for the purpose of comparing predicted functional category differences in accessory genome, COG functional group frequency distributions.

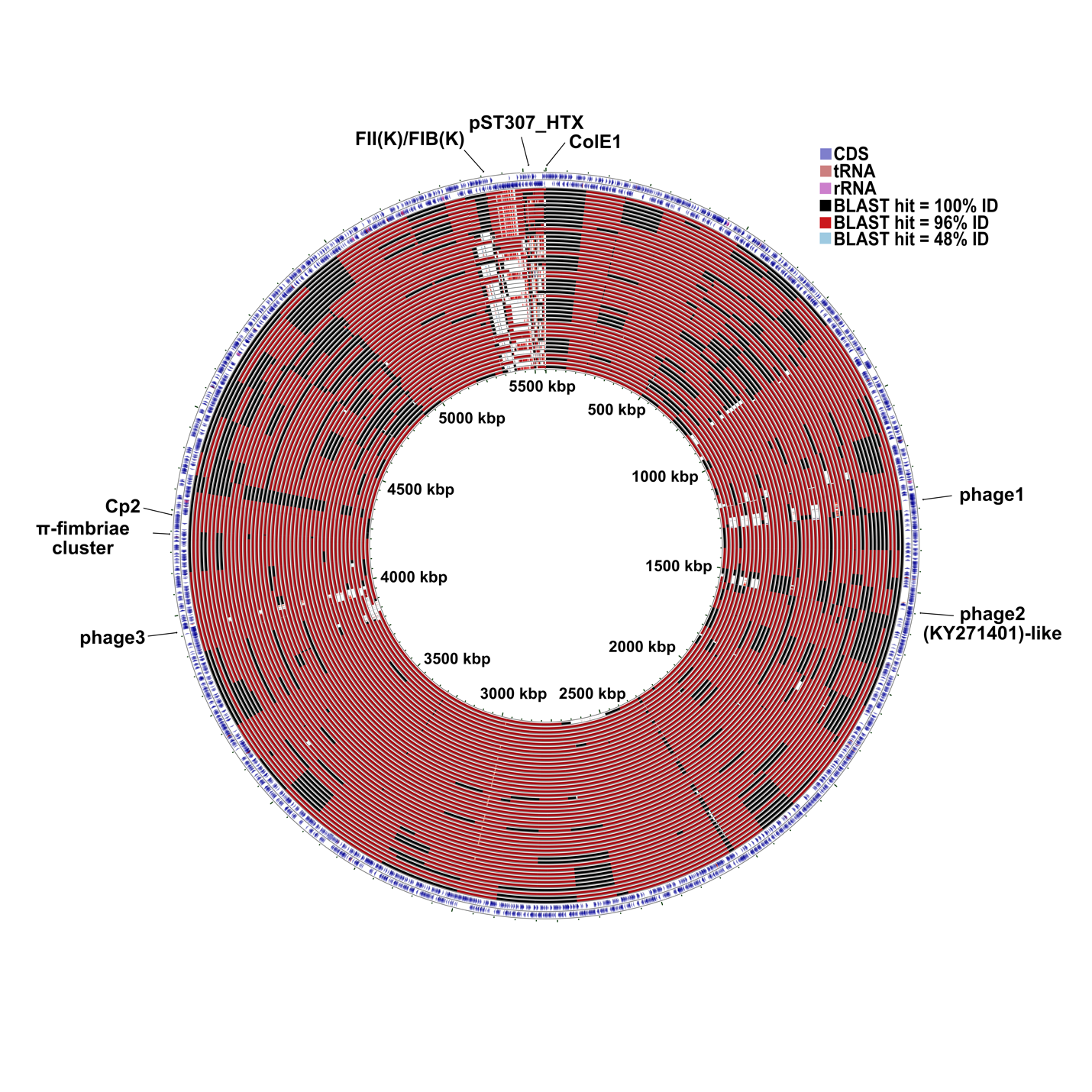

**Fig. S5.** Whole genome comparisons of all ST307 isolates in full cohort. Accessory genome features are labelled respectively. Isolate C234 was used for as reference (*i.e.* subject) for blastn results of DNA vs DNA comparisons. The noted differences in CG307 arise largely from differences in phage insertions as well as plasmid content as shown above. The well conserved type-1 fimbriae cluster as well as the unique capsule synthesis (Cp2) are labelled accordingly.

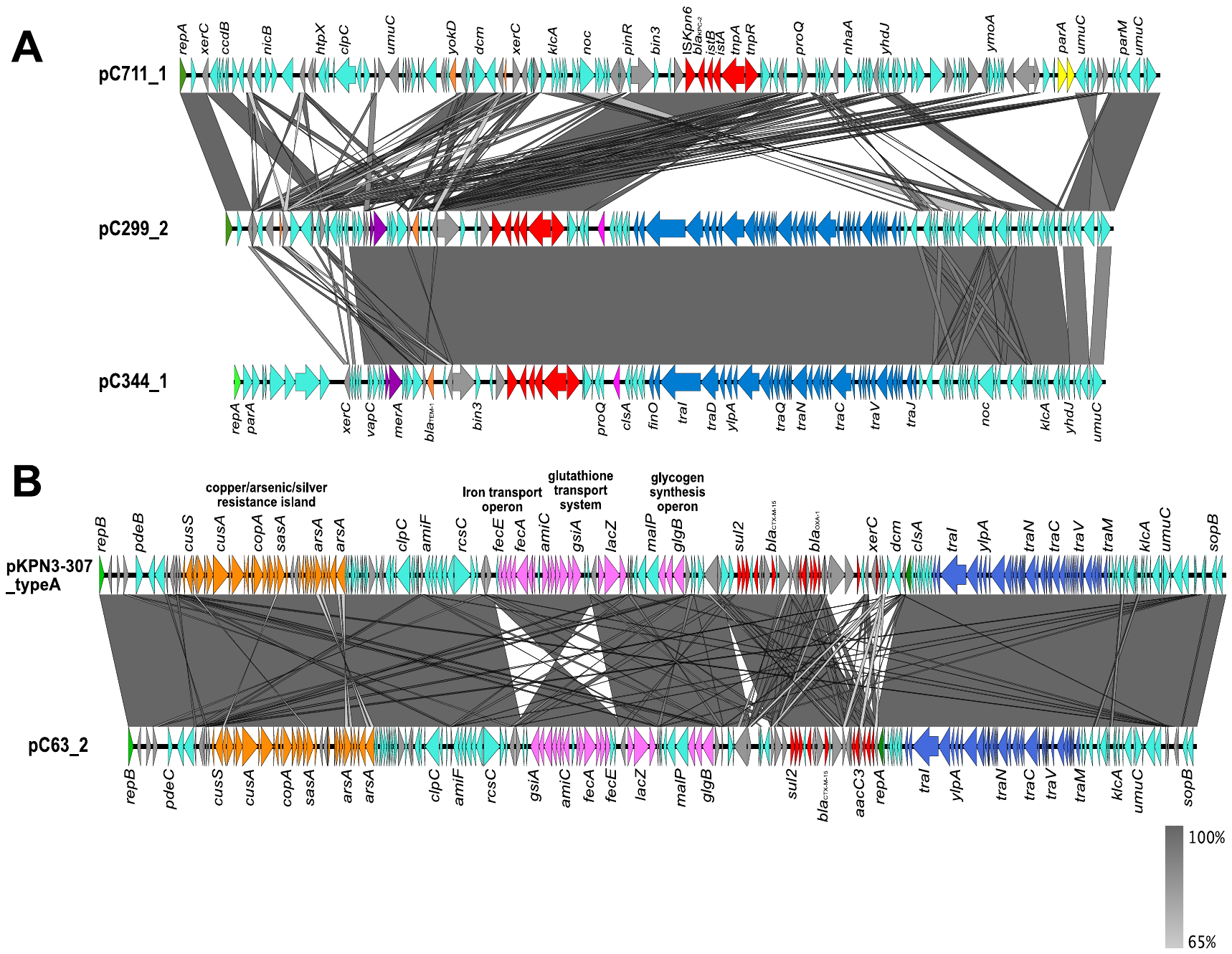

**Fig. S6.** Alignment of multi-replicon type F plasmids. **(A)** Three F-type plasmids with predicted conjugative operon genes (pC299_2 [CG307] and pC344_1 [Cg258]) and a type R plasmid that shares novel Rep-3 family *repA* gene (dark green) with pC299_2 (pCG307_HTX). Coding sequences in red denote Tn*4401a*. The ~34 Kbp *tra* operon is denoted in blue. pKpQIL rep-3 *repA* (light green), FIIK *repA* (pink), R-plasmid signatures (yellow), AMR genes (orange), mercury resistance operon (purple), MGEs (grey), Other CDS (turquoise) labelled accordingly. **(B)** pKPN3-307_typeA and pC763_2 (CG307) alignment that share 99.99% blastn identity and 93% coverage. *repB* gene is shaded green. Heavy metal resistance determinants (orange), virulence determinants (pink), resistance determinants (red), MGEs (grey), other coding DNA sequences (turquoise), and *tra* operon (blue) shaded accordingly.

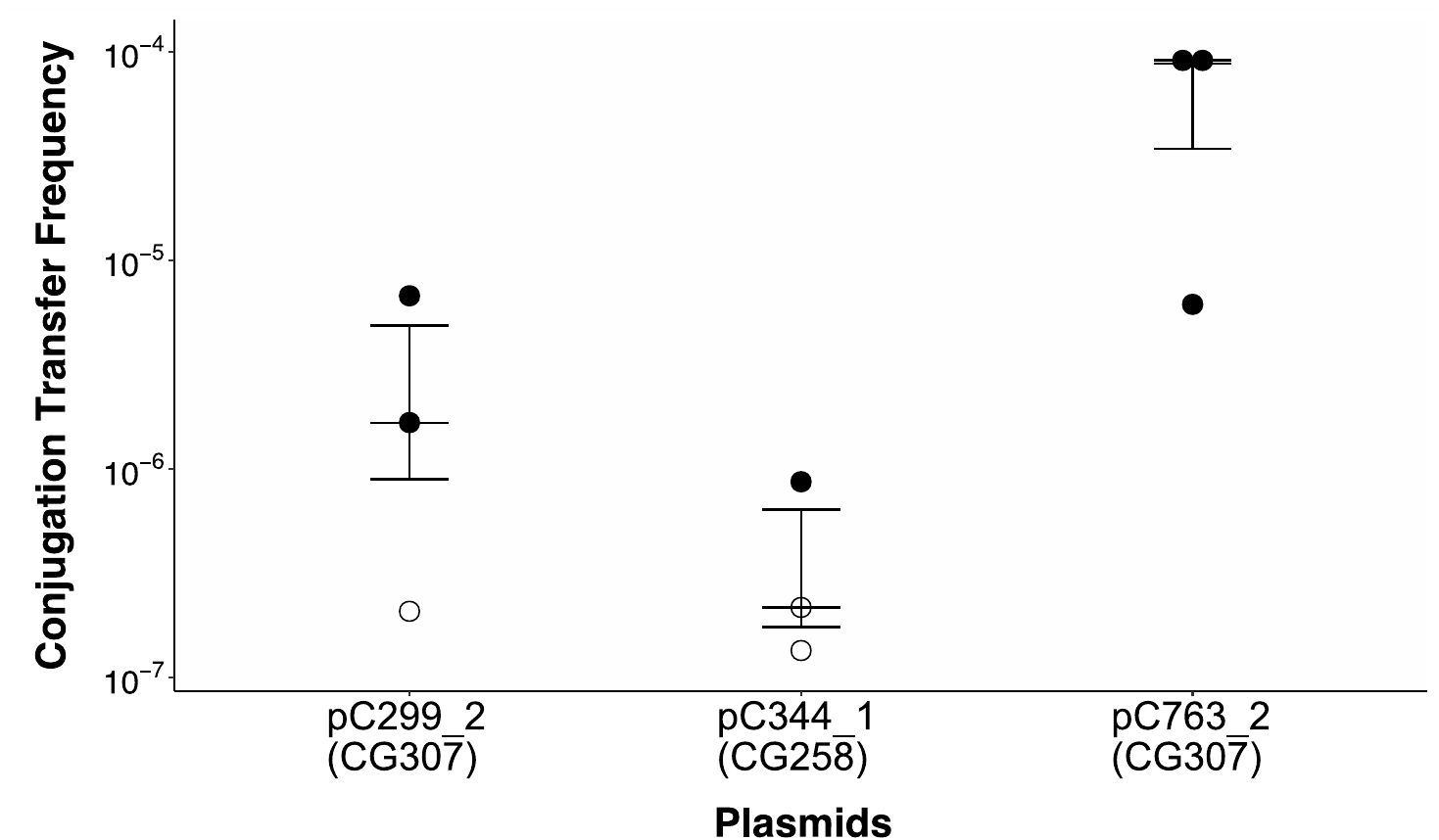

**Fig. S7.** Conjugation transfer frequency from predicted conjugative CG307 and CG258 plasmids. Conjugation transfer frequency calculated by transconjugant frequency (CFU/mL)/donor frequency (CFU/mL). pC992_2 – pCG307_HTX; pC344_1 – pKpQIL; pC763_2 – pKPN3_ST307_typeA-like plasmid. Each circle indicates the conjugation transfer frequency per experiment. Open circles indicate no transconjugants detected above the limit of detection. The middle bar indicates the median conjugation transfer frequency with bars equal to the standard error. One-way ANOVA p-value = 0.06.
